# Analysis of common PI3K-AKT-MTOR mutations in pediatric surgical epilepsy by droplet digital PCR reveals novel clinical and molecular insights

**DOI:** 10.1101/2021.06.09.21257462

**Authors:** Filomena Pirozzi, Matthew Berkseth, Rylee Shear, Lorenzo Gonzalez, Andrew E. Timms, Josef Sulc, Emily Pao, Nora Oyama, Francesca Forzano, Valerio Conti, Renzo Guerrini, Emily S. Doherty, Sulagna C. Saitta, William B. Dobyns, Edward Novotny, Jason N.N. Wright, Russell P. Saneto, Seth Friedman, Jason Hauptman, Jeffrey Ojemann, Raj P. Kapur, Ghayda M. Mirzaa

## Abstract

Focal malformations of cortical development (FMCD) including focal cortical dysplasia (FCD), hemimegalencephaly (HMEG) and megalencephaly (MEG), constitute a spectrum of neurodevelopmental disorders associated with brain overgrowth, cellular and architectural dysplasia, intractable epilepsy, autism, and intellectual disability. Importantly, FCD is the most common cause of intractable pediatric focal epilepsy. Gain and loss of function mutations in the PI3K-AKT-MTOR pathway have been identified in this spectrum, with variable levels of mosaicism and tissue distribution. In this study, we aimed to assess droplet digital Polymerase Chain Reaction (ddPCR) as a first-tier molecular diagnostic method, as well as define genotype-phenotype relationships among the most common PI3K-AKT-MTOR pathway mutations in FMCD.

A total of 144 specimens, including 113 brain samples, were collected from 58 individuals with intractable focal epilepsy phenotypes including FCD, MEG, HMEG and other types of developmental cortical lesions. We designed an ultra-deep and highly sensitive molecular diagnostic panel using ddPCR for six of the most common mutations in three PI3K-AKT-MTOR pathway genes, namely *PIK3CA* (p.E542K, p.E545K, p.H1047R), *AKT3* (p.E17K) and *MTOR* (p.S2215F, p.S2215Y). We quantified the level of mosaicism across all samples and correlated genotypes with key phenotype, neuroimaging and neuropathological data.

Pathogenic variants were identified in 17 individuals, with an overall molecular solve rate of %. Variant allele fractions (VAF) ranged from 0.1% to 22.67% across all positive samples. Our data shows that *MTOR* mutations are mostly associated with FCD, whereas *PIK3CA* mutations are more frequent in the HMEG-DMEG spectrum. The presence of one of these common PI3K-AKT-MTOR-mutations correlated with earlier onset of seizures. However, levels of mosaicism did not correlate with the severity of the cortical malformation by neuroimaging or neuropathological examination. Interestingly, we could not identify the six most common pathogenic variants in other types of cortical lesions (e.g., polymicrogyria or mesial temporal sclerosis) suggesting that PI3K-AKT-MTOR mutations are specifically causal in the FCD-HMEG-MEG spectrum. Finally, our data suggest that ultra-deep targeted molecular analysis for the most common PI3K-AKT-MTOR mutations via ddPCR is an effective molecular diagnostic approach for FMCD phenotypes with a good diagnostic yield when paired with neuroimaging and neuropathology evaluations. The high sensitivity and low DNA input requirements suggests that ddPCR is an effective molecular diagnostic tool for disorders caused by somatic mutations with a narrow mutational spectrum, including specific subtypes of pediatric epilepsy surgical phenotypes such as FCD and HMEG.

## Introduction

Focal malformations of cortical development (FMCD) including focal cortical dysplasia (FCD), hemimegalencephaly (HMEG) and dysplastic megalencephaly (MEG), constitute a spectrum of developmental brain disorders associated with brain overgrowth, cellular dysplasia, and co-morbidities such as intractable epilepsy, autism, and intellectual disability. Predominantly somatic or mosaic gain of function (GOF) mutations in the PI3K-AKT-MTOR pathway have been identified in this spectrum.^1–8^ Establishing a molecular diagnosis for these disorders has significant benefits for affected families, including understanding the natural history, informed genetic counseling, as well as more personalized therapeutic interventions. While traditional molecular diagnostic approaches, such as Sanger sequencing or standard-depth multi-gene panel or exome sequencing using peripheral blood-derived DNA, reliably detect inherited or germline genetic variants, these methods lack the sensitivity to detect low-level postzygotic (mosaic) mutations. Further, the high genomic input requirements for these tests renders them impractical in this spectrum due to paucity of affected (or lesional) tissues such as surgical brain samples. To address these challenges, we designed an ultra-deep targeted sequencing panel using droplet digital polymerase chain reaction (ddPCR) that includes six of the most common PI3K-AKT-MTOR mutations identified in this spectrum.^9,10^

Droplet digital PCR (ddPCR) is an allele-specific detection method where the PCR reaction is partitioned into ∼20,000 separate emulsion droplets. This method has been optimized to quantify extremely low-frequency single nucleotide variants and copy number abnormalities (down to 0.01% variant allele fraction; VAF).^11^ Therefore, ddPCR has been increasingly used as a clinical diagnostic method to efficiently and reliably detect and quantify common cancer-associated variants (such as the common *PIK3CA* hotspot mutations – p.C420R, p.E542K, p.E545K, p.H1047L, and p.H1047R – for *PIK3CA*-related overgrowth disorders (PROS), and the *BRAF* V600E mutation for brain and thyroid tumors, among others), and has also been recently used in other pediatric overgrowth, vascular and lymphatic malformation phentypes.^12–, 15^ The PI3K-AKT-MTOR-related developmental brain disorders, including most notably FCD and HMEG, have a narrow mutational spectrum, with a few “hotspot” mutations or variants representing a sizable portion of molecularly-solved cases.^4, 6, 9, 10^ Therefore, we tested these common mutations as a first tier molecular method to complement the neuroimaging and neuropathology diagnosis, in a large cohort of affected individuals to efficiently identify mutations and studying the relationship between levels of mosaicism and key neuroimaging and neuropathological features.

## Materials and methods

### Human Subjects

Patients diagnosed with focal intractable epilepsy requiring epilepsy surgery were prospectively enrolled in the Developmental Brain Disorders Research Program under an IRB approved protocol at Seattle Children’s Hospital (IRB#13291). All patients signed an informed consent according to the Declaration of Helsinki or were de-identified and included in the study under the IRB approved “waiver of consent”. Brain tissue samples were collected during clinically indicated epilepsy surgery. Additional samples (such as saliva, peripheral blood, or skin fibroblasts) were also collected from affected individuals, when possible.

### Inclusion and exclusion criteria

Individuals with epilepsy who underwent epilepsy surgery and confirmed by neuroimaging and/or neuropathologic examination to have one of the following diagnoses were included in the study: FCD, HMEG, DMEG, malformations of cortical development, or other developmental lesions. Patients with brain tumors were excluded from this study. Patients with predominantly somatic phenotypes (such as overgrowth, vascular or lymphatic malformations) who did not have any central nervous system involvement (i.e., cortical malformations or megalencephaly) were excluded from this study.

### Neuroimaging

All patients evaluated at the Pediatric Epilepsy Surgery center at Seattle Children’s Hospital had brain magnetic resonance imaging (MRI) using the Siemens 1.5T (Symphony or Avanto) or 3T (TrioTim or PRISMA) systems, or external site data of equivalent quality. Common pre-operative structural data included sagittal T1 MPRAGE (isotropic 1mm), axial and coronal T2 (2-4mm), and axial fluid-attenuated inversion recovery (FLAIR) images (2-4mm). We performed diffusion weighted images skull stripping with the brain extraction tool in FSL on T1-data then made 3-dimensional renderings with BioImage Suite.^16^ Where indicated as helpful for lesion evaluation, the same skull stripped mask was used for the T2- or FLAIR-data. Coordinates for brain tissue samples were created using the image guided surgery system (Metronic Stealth) with intra-operative x-y-z coordinates downloaded and then transformed into MRI space and overlaid using BioImage Suite. Brain MRIs were examined and assessed for the type, distribution and severity of the cortical dysplasia, as well as the presence of any other notable brain MRI abnormalities. All brain images were assessed for laterality of findings (i.e., unilateral or bilateral cortical malformations), symmetry, gradient (anterior vs. posterior), lobes or brain regions affected, white matter abnormalities and any mass effect.

### Neuropathological examination

Histology sections of all resected brain tissues underwent comprehensive examination to establish a neuropathologic diagnosis. Hematoxylin-and-eosin (H&E)-stained sections were examined from formalin-fixed, paraffin-embedded (FFPE) from each site of brain tissue resection. In addition, immunohistochemistry was performed on sections from at least one and often multiple FFPE tissue blocks by established methods using the mouse monoclonal anti-NeuN (Millipore, MAB 377) and mouse monoclonal anti-GFAP (Dako, Mo761) antibodies to assess neuronal and glial cell populations. Histopathologic FCD was diagnosed and subtyped based on criteria established by the International League Against Epilepsy Task Force and described in detail by Blumcke and colleagues.^17, 18^ For patients with histologically confirmed FCD type 2a or 2b, tissue sections from multiple brain regions, including areas with no histopathologic dysplasia, underwent additional immunohistochemical staining using the rabbit polyclonal anti-phospho-S6 (pS6) ribosomal protein (Cell Signaling 2211). The percentage of pS6-positive cortical gray matter in a given site was estimated based on the fraction of cortical neurons with moderate-to-markedly intense cytoplasmic pS6 immunoreactivity. All of the neuropathological assessment, including pS6 immunolabeling assessment, was performed by a board-certified pediatric pathologist (RPK), who was “blinded” with respect to the neuroimaging and ddPCR data.

### Sample processing for ddPCR

We collected 144 specimens from a cohort of 58 children diagnosed with FCD, MEG, HMEG, DMEG, and other malformations of cortical development (MCD) including polymicrogyria (PMG) and multifocal cortical dysplasia. Samples included fresh frozen (*n*=101) and formalin-fixed paraffin-embedded (FFPE, *n*=12) brain tissue from epilepsy surgeries (total brain samples *n*=113), saliva (*n*=24), peripheral blood (*n*=3), skin (*n*=2), and buccal swab (*n*=2). All brain samples were either snap-frozen tissue located immediately adjacent to tissues fixed and embedded for neuropathologic examination, or were sections from FFPE tissue blocks used for neuropathology. Genomic DNA extraction was performed using standard protocols with PureLink™ Genomic DNA Mini Kit (Invitrogen, cat. num. K1820-01). FFPE samples were processed using the GeneRead DNA FFPE Kit (Qiagen, cat. num. 180134) following the manufacturer’s protocol. DNA quality and concentration were quantified using the NanoDrop 3300 (ThermoFisher Scientific), and the Qubit 4 Fluorometer (ThermoFisher Scientific). If DNA quality was suboptimal (defined by 260/280 ratios below 1.8 and/or 260/230 ratio below 2), samples were cleaned and concentrated using a column-based kit (Zymo research Genomic clean and concentrator, cat. num. D4011) and re-quantified before proceeding with ddPCR.

### Droplet digital polymerase chain reaction (ddPCR) testing

A custom ddPCR-based panel was designed using probes targeting six common mutations in three PI3K-AKT-MTOR pathway genes, namely *PIK3CA* (p.E542K, p.E545K, p.H1047R), *AKT3* (p.E17K), and *MTOR* (p.S2215Y, p.S2215F). We selected these specific variants as they are the most common variants seen in FCD-HMEG phenotypes,^10, 19, 20^ as well as the most common mutations in these genes in somatic tissues in cancer (COSMIC, Catalogue of Somatic Mutations in Cancer). ddPCR probes were purchased as validated assays from Bio-Rad (see Supplementary Table 1 for list of probes used). As FFPE samples can have high levels of DNA degradation and nonreproducible sequence artifacts, especially C:G > T:A transitions that can increase the chance of false positive results, all FFPE derived-DNA samples were treated with uracil DNA glycosylase (UDG) to eliminate PCR amplification carryover prior to ddPCR testing. This method has been successfully used to detect mosaic cancer-associated mutations in FFPE samples.^21, 22^ Approximately 8-10 ng of DNA was used per ddPCR reaction, and all samples were tested in technical quadruplicate using previously published methods.^12, 23^ Briefly, the ddPCR reaction mix was assembled from 10 μL ddPCR Supermix for Probes (Bio-Rad, 1863024), 1 μL ddPCR validated mutant assay (Bio-Rad, see **Supplementary Table 1** for full list), and 12 μL water per reaction. A restriction enzyme was not included due to the low DNA input. 20 μL of the ddPCR mastermix along with 70 μL Droplet Generation Oil were loaded into a disposable droplet generator cartridge and placed into a QX200 Droplet Generator (Bio-Rad). Generated droplets were transferred to a 96 well PCR plate and placed in a thermal cycler for amplification (see Supplementary Table 1 for probe-specific annealing temperature). Amplification cycles were set as follows: denaturing phase at 95°C for 10 minutes, followed by 40 cycles of 94°C for 30 seconds and annealing/extension temperature for 1 minute, with a final denaturation at 98°C for 10 minutes and hold at 4°C. ddPCR plates were then placed into a QX200 Droplet Reader (Bio-Rad) for fluorescent quantification. Data analysis was performed using the Quantasoft AP software. Positive fluorescence thresholds for mutant (FAM) and wild-type (HEX) associated probes were established using three controls included in quadruplicate in each run: a no template control (NTC), a wild-type (non-mutant) control, and a mutation positive control (with VAF ranging from 3-30%). Data from quadruplicate wells were pooled for each sample and VAF% calculated. To account for rare non-specific mutant-positive (i.e., false positive) droplets, the calculated VAF for each sample was compared to the wild-type control run in the same plate. Delta Confidence Intervals (DCI) were calculated by subtracting the Poisson-normalized maximum fractional abundance (95% CI) for the wild-type control from the Poisson-normalized minimum fractional abundance (95% CI) for each sample. Due to limited sample quality and amount from some samples, such as FFPE-derived brain samples, we used more stringent customized cut-offs to minimize false positive results where samples were deemed positive for a given mutant allele when their DCI exceeded 0.045 (rather than the DCI cut-off of 0 per the Bio-rad analysis protocol) and the average number of wild-type positive droplets for the pooled set of quadruplicate wells exceeded 3000 droplets. If the number of wild-type droplets was between 500 and 3000, samples were retained in the analysis only if the DCI was above 1 and/or other samples from the same patient showed a clearly positive result. These cut-offs were empirically established by analyzing borderline-positive samples (with a DCI between 0 and 0.05) that were assayed in multiple independent experiments with different DNA concentrations. Examples of negative and positive samples Quantasoft 2D plots are shown in **Supplementary Fig. 1B**. When samples were analyzed in more than one experiment, the run producing the highest wild-type positive droplet count and highest DCI was reported here.

### Statistical analysis

Statistical analysis and relative graph generation was performed in GraphPad Prism version 9.0.2. The appropriate statistical test was chosen based on the number of replicates and type of comparison of interest. Details on specific statistical tests and relative results are provided with the associated data below.

### Data availability

All data generated or analysed during this study are included in this published article and its supplementary information files. The raw data generated during and/or analysed during the current study are available from the corresponding author on request.

## Results

We screened a cohort of 58 children who underwent epilepsy surgery and sub-grouped them based on their neuroimaging and neuropathologic diagnoses to four groups: FCD (*n*=33), HMEG-DMEG (*n*=11), other MCD (*n*=7), or other cortical lesions (*n*=7). The latter category includes gliosis (*n*=3), hypoxic ischemic encephalopathy (HIE, *n*=1), meningoangiomatosis (*n*=1), mesial temporal sclerosis (MTS, *n*=1) and stroke (*n*=1). Notably, one individual with HMEG (LR16-242) presented contralateral hemimicrencephaly. This is not the first case of HMEG with contralateral hemimicrencephaly, suggesting that the non-HMEG hemisphere might be at risk of developmental delay as well, although studies on the contralateral hemisphere are usually limited in these patients.^24, 25^ A summary of the samples and distribution of the cohort, as well as representative brain MR images, are shown in **Fig. 1**. Of the 58 children tested, 43 had at least one brain sample collected, with 7 individuals having more than 5 brain regions analyzed each (**Fig. 2A**). Following mutation detection, as determined by the cutoff criteria (stated above, **Supplementary Fig. 1B** for representative positive and negative 2D plots results), additional peripheral samples from those affected individuals were serially tested to further discern the levels of mosaicism (VAF) across all available tissues. The 6 hotspot mutations (*PIK3CA* p.E542K, p.E545K, p.H1047R; *MTOR* p.S2215F and p.S2215Y; *AKT3* p.E17K) were detected in 42 samples (*n*=6 saliva, *n*=36 brain) belonging to 17 individuals (29.31% of total cohort); with VAF ranging from 0.16% to 22.67%, as summarized in **Table 1**. The overall yield for molecular diagnostic testing, or solve rate, was 29.31%, albeit dependent on the number and types of tissues available per patient. We were able to identify one of these 6 hotspots in 13.33% of individuals on whom no brain tissues were available (2/15 individuals), and 34.88% among patients with brain samples (15/43 patients). Specifically, we detected one of the 6 mutations in 20% of individuals with one brain tissue tested (6/30 individuals), increasing to 53.85% solve rate for individuals with at least two brain specimens (range 2-15 brain specimens per patient, 7/13 children), in contrast with the 25% solve rate for patients with saliva samples (with or without paired brain samples, 6/24 patients) (**Fig. 2B**, **Supplementary Fig. 1A**). Interestingly, these hotspot mutations were only detectable in brain or, much more rarely, saliva samples, but not in peripheral blood or skin derived samples, albeit all non-brain samples were much fewer in this cohort.

**Figure 1:**
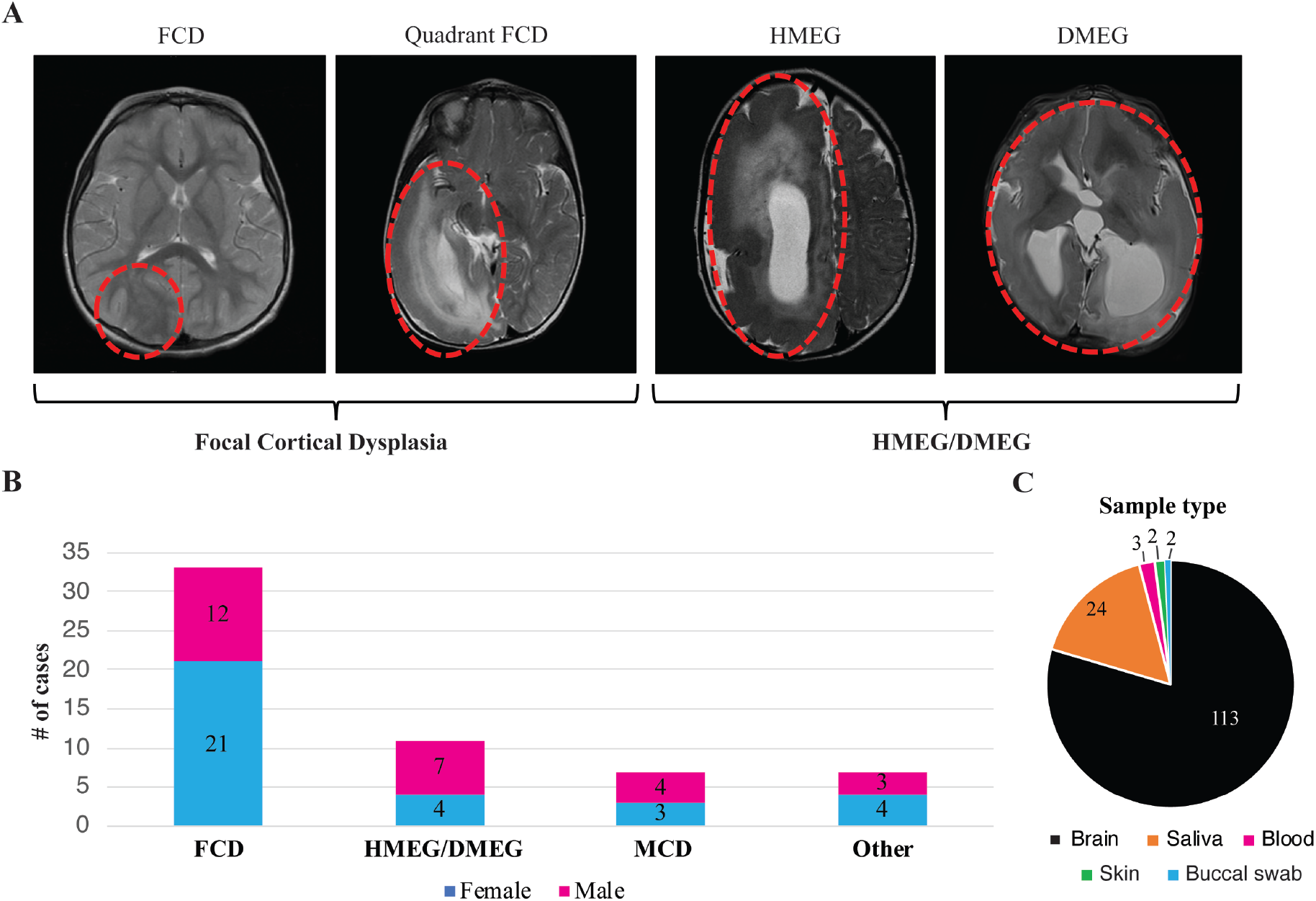
Overview of the cohort and samples. **(A)** Representative brain MRIs of patients with focal cortical dysplasia (FCD), hemimegalencephaly (HMEG), and dysplastic megalencephaly (DMEG). Representative affected brain areas are highlighted by red ovals. Patients diagnosed with FCD and quadrantic dysplasia were clustered in the FCD category, while patients diagnosed with HMEG or DMEG were clustered in the HMEG/DMEG category. **(B)** Bar graph representing the cohort distribution stratified by the 4 clinical diagnosis categories (FCD, HMKEG/DMEG, and malformations of cortical development, MCD, as well as other diagnoses) and sex. The “other” diagnoses include 7 cases with gliosis (n=3), hypoxic ischemic encephalopathy (HIE, n=1), meningoangiomatosis (n=1), mesotemporal sclerosis (MTS, n=1) and stroke (n=1). **(C)** Pie chart representing types and numbers of samples collected from the 58 individuals in this series. The vast majority of samples were brain (N = 114, fresh frozen or FFPE).

**Figure 2:**
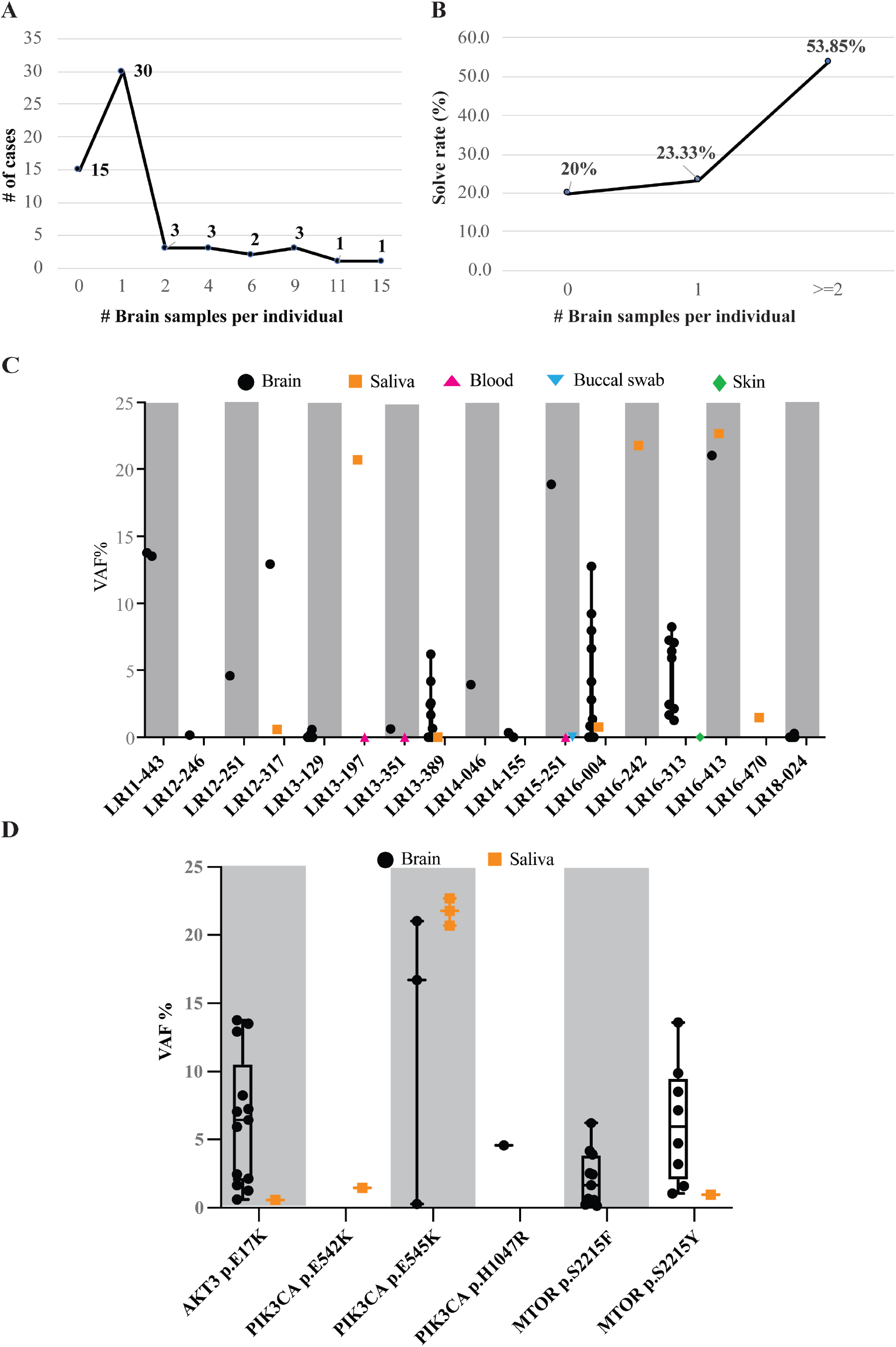
Molecular results. **(A)** Number of brain samples per patient in our cohort. Most of the patients had at least 1 brain sample (N = 30). **(B)** The molecular yield (or solve rate) stratified by number of brain samples. The solve rate was calculated as number of positive cases with a given number of brain samples over the total number of cases with the same number of brain samples, and expressed as a percentage. As expected, our solve rate was increased with increasing numbers of brain samples, indicating that the number and type of samples per patient is crucial to identify a causal mosaic mutation. **(C)** Variant allele fraction (VAF%) for the 17 mutation-positive patients. The y-axis represents the VAF%, while patients are listed on the x-axis. All samples belonging to the 17 patients are represented, including negative ones (crossing the 0 on the y-axis). Gray shaded boxes are used to represent every other patient to delineate samples belonging to each individual. Different tissues are represented as indicated in the graph legend. Only brain and saliva samples were mutation positive, whereas blood, buccal swab and skin fibroblast-derived samples were negative. **(D)** Positive samples stratified by the PI3K-AKT-MTOR hotspot mutation to visualize the range of VAF for each mutation across individuals. Notably, the PIK3CA, p.E545K, variant had the widest VAF% range and was the only hotspot to be present at higher VAF% in saliva rather than brain samples.

**Table 1:** Summary of mutation positive patients. List of the diagnoses, clinical features and molecular results for the 17 mutation positive cases. When a dash (-) is present in the “other clinical features cells, it refers to absence of other clinical features. Abbreviations: DX: diagnosis; MRI: Magnetic Resonance Imaging; NP: neuropathology; VAF: Variant Allele Fraction; HMEG: hemimegalencephaly; FCD: focal cortical dysplasia; DMEG: dysplastic megalencephaly; N/A: not available.

| Patient ID | DX (MRI) | DX (NP) | Seizure – age at onset | # epilepsy surgeries | Other clinical features | Pathogenic variant | VAF% range | VAF% Average | # samples | # brain samples | # positive samples |
| --- | --- | --- | --- | --- | --- | --- | --- | --- | --- | --- | --- |
| LR12-246 | HMEG | FCD 2a | 3 days | 2 | Sacral lipoma | MTOR (p.S2215F) | 0.16 | 0.16 | 1 | 1 | 1 |
| LR13-129 | FCD | FCD 2b | 11 months | 4 | – |  | 0.24-0.57 | 0.13 | 9 | 9 | 2 |
| LR13-389 | FCD | FCD 2a | 5 weeks | 2 | Amblyopia of right eye, dissociated vertical deviation, anisometropia |  | 0.65-6.22 | 1.14 | 16 | 15 | 6 |
| LR14-046 | FCD | FCD 2b | 7 months | 2 | – |  | 3.91 | 3.91 | 1 | 1 | 1 |
| LR14-155 | FCD | FCD 2b | 5 years | 1 | – |  | 0.34 | 0.2 | 2 | 2 | 1 |
| LR13-197 | DMEG | FCD 1b | 29 days | N/A | Right lower extremity hemihypertrophy with left-sided facial asymmetry | PIK3CA (p.E545K) | 20.68 | 10.36 | 2 | 0 | 1 |
| LR15-251 | HMEG | N/A | 1 day | 3 | Flat feet, translucent skin, sebaceous nevus on left cheek, congenital recurrent nocturnal apneas, adenoidectomy |  | 18.86 | 6.33 | 3 | 1 | 1 |
| LR16-242 | HMEG | N/A | 1 day | No | Enlarged R cheek and eye (ipsilateral to HMEG), swirled brown macule on R cheek extending to neck |  | 21.76 | 21.76 | 1 | 0 | 1 |
| LR16-413 | HMEG | FCD | 2 days | 1 | Hypertrophic left lip/cheek |  | 21.01-22.67 | 21.84 | 2 | 1 | 2 |
| LR18-024 | FCD | FCD 2a | 6 years | 2 | – |  | 0.28 | 0.11 | 9 | 9 | 1 |
| LR11-443 | HMEG | FCD 2a | 3 days | 2 | Vascular malformations of the skin, organomegaly, scoliosis, visual loss in left eye | AKT3 (p.E17K) | 13.51-13.76 | 13.64 | 2 | 2 | 2 |
| LR12-317 | HMEG | N/A | 7 days | 1 | Vascular abnormalities of lower extremities; PWS on the legs, two small lesion on chest and arms |  | 0.58-12.93 | 6.76 | 2 | 1 | 2 |
| LR13-351 | FCD | FCD 2a | ~1 month | – | – |  | 0.62 | 0.32 | 2 | 1 | 1 |
| LR16-313 | FCD | FCD 2a | 4 days | 1 | – |  | 1.26-8.25 | 4.25 | 10 | 9 | 9 |
| LR16-004 | HMEG | FCD 2a | 3 weeks | 1 | – | MTOR (p.S2215Y) | 0.96-13.59 | 4.23 | 12 | 11 | 9 |
| LR16-470 | HMEG | N/A | N/A | N/A | Hyperpigmentation on several areas of the body | PIK3CA (p.E542K) | 1.47 | 1.47 | 1 | 0 | 1 |
| LR12-251 | FCD | FCD 2a | 3 years | 2 | – | PIK3CA (p.H1047R) | 4.58 | 4.58 | 1 | 1 | 1 |

### Molecular results

Using our ultra-deep and highly targeted ddPCR PI3K-AKT-MTOR panel, we were able to molecularly solve 17 cases, with 6 individuals positive for the *PIK3CA* p.E545K mutation; 5 positive for *MTOR* p.S2215F, 3 positive for *AKT3* p.E17K, and one positive patient for each of the following mutations: *PIK3CA* p.E542K, *PIK3CA* p.H1047R, and *MTOR* p.S2215Y. The range of VAF for all 17 positive patients is graphically shown in **Fig. 2C**, including both positive and negative samples from these individuals. Individual LR16-413 had the highest overall VAF of the cohort, with two samples collected and positive for *PIK3CA* p.E545K mutation at 21.01% in frozen brain tissue and 22.67% in saliva. Interestingly, blood and fibroblast-derived DNA from this patient were previously tested by a single molecular inversion probe (smMIPs) targeted multi-gene panel of PI3K-AKT-MTOR pathway genes with no mutation detected.^4^ In contrast, individual LR12-246 had the lowest level of mosaicism (*MTOR* p.S2215F at VAF 0.16%) of the entire cohort in their only available sample (brain). Of note, when both brain and saliva-derived DNA samples (i.e., paired samples) were available for a given individual, the *PIK3CA* p.E545K mutation was the only one that had a higher VAF in saliva compared to brain (individual LR16-413) (**Fig. 2D**). The distribution of mutations and their VAF per sample for all mutation positive patients is shown in **Supplementary Table 2**. Patients who were tested for all 6 hotspots and were mutation-negative, as defined by the above-stated cutoff criteria, are listed in **Supplementary Table 3** (*n*=41).

### Genotype-phenotype correlation

To discern genotype-phenotype correlations among individuals with these common mutations, we stratified the mutation distribution based on the neuroimaging diagnosis and neuropathological findings. We found that 81.81.% of individuals clinically diagnosed with HMEG/DMEG had mutations in these 6 hotspots, compared to only 21.21% of FCD diagnoses by neuroimaging (**Fig. 3A**). While the number of positive patients within each category is not sufficient to establish a statistically significant correlation, we did observe a trend of association between an FCD diagnosis (by neuroimaging) and *MTOR* p.S2215F mutation (4 of the 7 positive patients), and between DEMG/HMEG diagnosis and *PIK3CA* mutations (p.E542K and p.E545K, 6 out of 9 positive cases), followed by *AKT3* p.E17K (3 of the 9 positive cases) (**Fig. 3A** and **B**). None of the cases diagnosed with MCD or other cortical lesions (*n*=14) were positive for the PI3K-AKT-MTOR mutations tested. We further stratified the cohort based on neuropathologic findings when available, and confirmed that FCD subtype 2a is more frequently caused by mutations in the PI3K-AKT-MTOR pathway, with a solve rate of 42.1% (**Fig. 3B**), a finding that has been corroborated by other studies. For 5 patients, we could only obtain pathology reports of FCD with no further specification of the FCD subtype (40% solve rate, **Fig. 3B** and **Supplementary Table 5**), while for 6 patients no neuropathology data was available (N/A in **Fig. 3B**, solve rate 66.66%). Interestingly, only one case out of ten diagnosed with FCD type 1 was positive for one of the hotspot mutations tested (patient LR13-197, *PIK3CA* p.E545K, 33.33% of FCD 1b cohort, **Fig. 3B**). Detailed ddPCR results for all positive patients are listed in **Supplementary Table 2**, more detailed clinical data for the 17 mutation positive patients are provided in **Supplementary Table 3**, and detailed ddPCR results for patients that were tested for the 6 hotspots and were negative (*n*=41) are listed in **Supplementary Table 4**.

**Figure 3:**
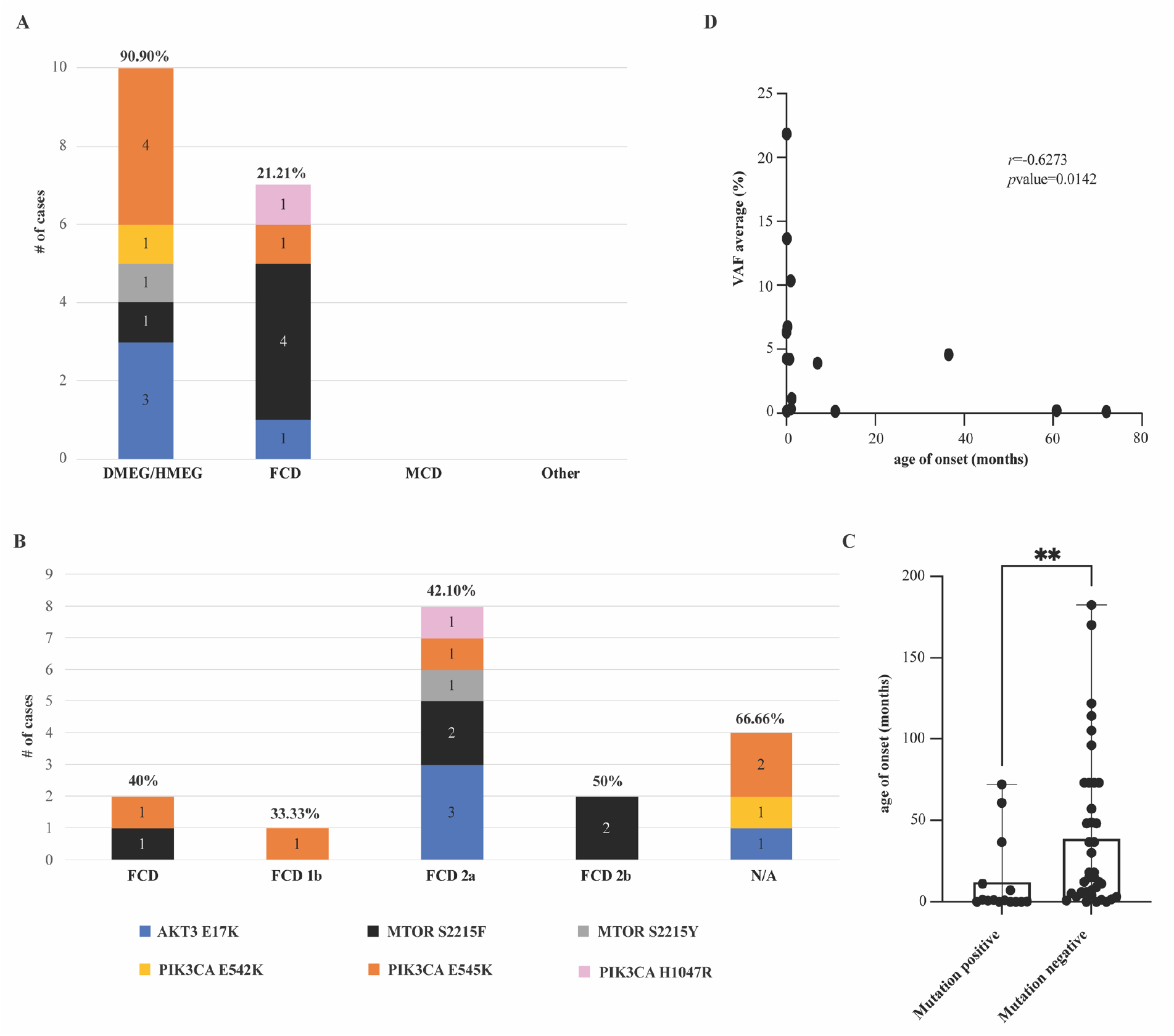
Genotype-phenotype correlations of PI3K-AKT-MTOR hotspot mutations. **(A)** Hotspot mutations detected in our cohort are stratified according to the four diagnostic categories and represented as stacked bar graphs. Notably, only FCD and HMEG/DMEG cases were positive for the 6 positive hotspots tested, while none of the MCD or “other” diagnoses were positive for these variants. The frequency of PIK3CA mutations was higher in DMEG/HMEG with a solve rate of 81.81%. In contrast, FCD had higher frequency of MTOR mutations (9/11 cases) with an overall solve rate of 24.24%. **(B)** The same data from graph A was stratified by the neuropathology classification/diagnosis and represented via stacked bar graph (number of cases in our cohort N=58). Solve rate for each category is shown as a percentage and calculated as number of positive cases/number of total cases within the same neuropathology subtype. FCD type 2a had the highest overall molecular yield in our series. **(C)** Bar graph showing the age of onset for seizure in the hotspot mutation positive (n=15) vs. hotspot mutation negative (*n*=39) cohorts. Each dot represents a data point, the top of the bar indicating the mean age of onset (12.79 months in mutation positive, 39.46 months in mutation negative) and error bars represent the full range. Kolmogorov-Smirnov test revealed a significant difference with \*\**p*value=0.0037. **(D)** Correlation analysis of seizure age of onset and VAF% in hotspot mutation positive patients (*n*=15). Spearman correlation analysis demonstrated a significant negative correlation with an *r*=-0.6873 and \*\**p*value=0.0042.

To determine whether these six hotspot mutations were associated with more severe epilepsy, we compared the age of seizure onset among the mutation positive and negative patients. Mutation positive patients had a significantly earlier age of onset for seizures (12.79 months, ±23.81, *n*=15) compared to mutation negative patients (average 39.46 months, ±47.72, *n*=39) using the Kolmogorov-Smirnov test (p-value=0.0037, **Fig. 3C**). Furthermore, we correlated the age of onset for seizures with the average VAF% for 15 of the positive patients, and discovered a negative correlation between these parameters, with earlier age of onset as the VAF% increases (*r*=0.6273, *p-*value=0.0142) using the Spearman correlation test (**Fig. 3D**, see **Table 1** for a complete list of seizure age of onset and VAF%).

To further assess whether there is a correlation between severity of phenotype and mutation burden in the brain tissues of interest, we analyzed the neuroimaging and neuropathology findings and paired them with VAF%, when matched samples were available. For 6 of the positive patients, we were able to overlay MRI imaging of the resection sites obtained during brain epilepsy surgery with their relative mutation burden (VAF%, **Fig. 4**). Notably, in several cases, the severity and distribution of the malformation by neuroimaging was much more extensive despite low VAF% in those regions. For example, patient LR13-129 (row 1 in **Fig. 4**) had extensive involvement of the frontal, temporal and parietal regions, and required a total of four epilepsy surgeries, and yet harbored the *MTOR*, p.S2215F, variant at extremely low VAF (0.24-0.57%). Similarly, patient LR18-024 had had extremely low VAF for *PIK3CA*, p.E545K, in only a few of surgically resected tissues despite much more extensive distribution by neuroimaging and neuropathology (row 4 in **Fig. 4**, **Fig. 5**). As a comparison, we assessed MRI images from patients that tested negative for all 6 probes in all their collected samples (**Supplementary Fig. 2**). However, we cannot rule out that these individuals do not harbor other genetic variants in the same genes or variants in other FMCD-associated genes limiting further correlations.

**Figure 4:**
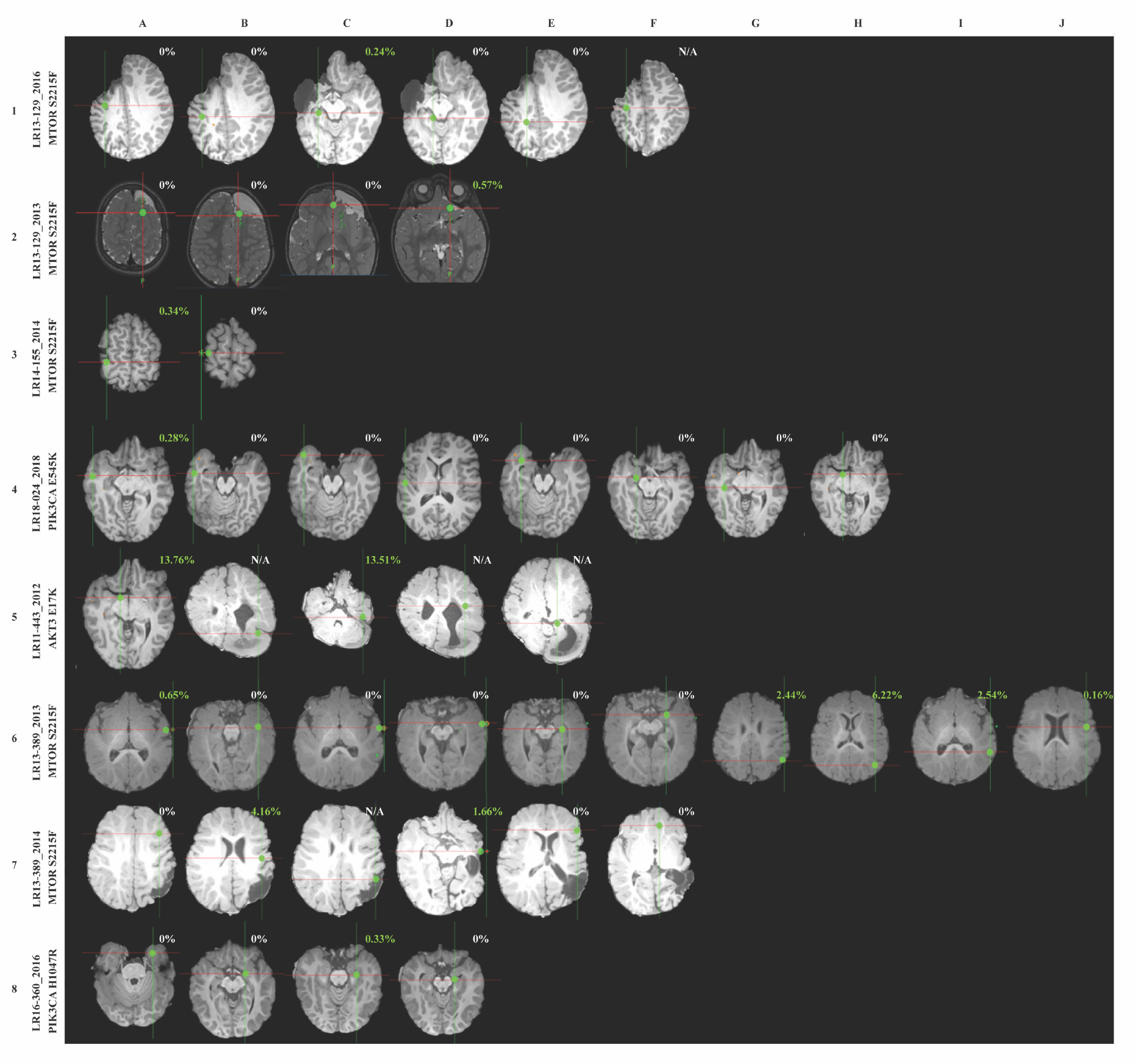
Neuroimaging findings and correlations with genotype. Representative brain MRIs showing brain biopsy or tissue resection locations alongside identified VAF of PI3K-AKT-MTOR hotspot mutations. For each image, cross-hairs reflect the site from which a tissue sample was obtained for molecular analysis, with letters at the top reflecting the multiple samples taken. At each location, ddPCR results are shown for the respective region. Green dots represent the exact location of resection, and ddPCR results are indicated as VAF% for positive samples. The year in which the surgery was performed is indicated after the underscore, as some patients underwent multiple brain surgeries (LR13-129, LR13-389). The reported VAF% did not directly correlate with the severity and distribution of visible cortical lesions.

**Figure 5:**
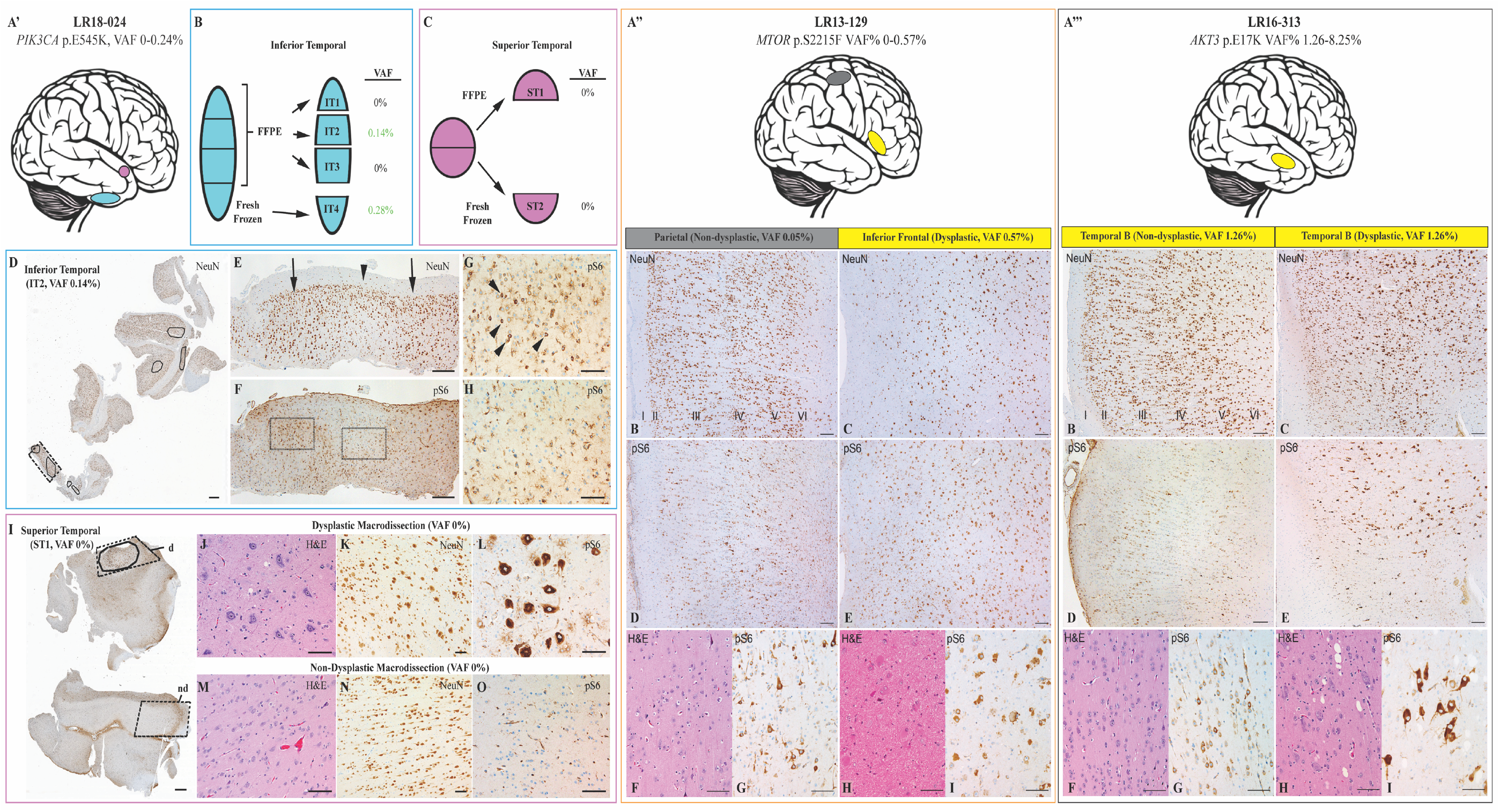
Histopathological findings and correlations with genotype in tissue samples from patient LR18-024, LR13-129 and LR16-313. Three representative individuals are depicted in the following panels: LR18-024 (panel A’ and following letters enclosed in light blue and magenta rectangles); LR13-129 (panel A” and following letters enclosed in the orange rectangle); LR16-313 (panel A”’ and following letters enclosed in black rectangle). **(A’)** Separate resected pieces of brain tissue (shown schematically) were received from the inferior (blue) and superior (pink) temporal lobe. (B) The inferior temporal piece was divided into 4 portions: 3 were formalin-fixed and embedded in paraffin (FFPE, IT-1, −2, −3) and one (IT4) was fresh frozen, as shown in the scheme. VAF% results obtained via ddPCR for the four portions are shown next to each sub-resection. (C) The superior temporal piece was divided in two halves: one half submitted as FFPE (ST1) and one half as fresh frozen (ST2). (D) A NeuN immunostained section at low magnification shows the entire block-face from IT-2 with areas of cortical dysplasia (outlined). (E) Higher magnification of the region indicated by dashed rectangle in D, which shows abnormal clustering of neurons in two areas of histological dysplasia (arrows) with intervening histologically normal cortex (arrowhead). (F) pS6 immunolabelling of a tissue section adjacent to (E), including dysplastic (G) and non-dysplastic (H) foci. pS6-immunoreactive neurons (arrowheads in F) are present in the dysplastic focus, whereas only glia cells show immunoreactivity in the non-dysplastic zone (G). (I) A pS6 immunostained section showing the entire block face from the superior temporal piece (ST1) with the dysplastic focus encircled. The dashed outlines indicate areas that were macrodissected from the block to enrich for histologically dysplastic (d) and non-dysplastic (nd) cortex. Images from the macrodissected regions highlight the cellular enlargement, disorganization, and strong pS6 immunoreactivity that differentiate the dysplastic (J-L) and nondysplastic (M-O) cortex. Scale bars: D,H – 1 mm; E,F – 300 μm; G,H, J-O – 100 μm. **A’’)** Schematic representation of the resection locations for patient LR13-129. Immunohistochemical and ddPCR findings in the histologically non-dysplastic parietal (B,D,F,G) and dysplastic (FCD type IIB) inferior frontal (C, E, H, I) resection specimens from patient LR13-129. (B) NeuN labeling of neurons in a low magnification field from the parietal resection specimen shows intact six-layered cortical gray matter as indicated by Roman numerals. The neurons appear cytologically normal (F), but a significant subset show strong perikaryal pS6 immunoreactivity (D,G). (C) In contrast, NeuN immunostaining shows marked disorganization of the cortical lamina in the inferior temporal region with balloon cells (H) and diffuse positive pS6 immunoreactivity in nearly the entire neuronal population (I). Scale bars: B-E, 200 μm; F-I, 100 μm. **(A’’’)** Schematic representation of the resection location for patient LR16-313. Immunohistochemical and ddPCR findings in the histologically non-dysplastic (B, D, F, G) and dysplastic area (C, E, H, I) from the temporal lobe resection (block B) LR16-313. (B) NeuN labeling of neurons in a low magnification field from a portion of the temporal lobe resection specimen shows intact six-layered cortical gray matter as indicated by Roman numerals. The neurons appear cytologically normal (F), despite having a detectable mosaic burden in this region (VAF 1.26%). In contrast, NeuN immunostaining shows disorganization of the cortical lamina in a portion of the same region (C) with atypical large neurons (H) and intense positive pS6 immunoreactivity in a subset of the neuronal population (I). Scale bars: B-E, 200 μm; F-I, 100 μm. Overview of ddPCR results for all samples belonging to these three individuals with matching neuropathology evaluation is presented in Supplementary Table 6, with the negative blocks (shaded in gray) and positive blocks (shaded in yellow) matching the ones represented in this figure.

Next, we aimed to identify correlations between genotypes and neuropathological features for all 58 patients in our cohort, as summarized in **Supplementary Table 5**. For 3 of the mutation-positive patients, we performed a matched analysis using brain samples from the same regions that were independently analyzed by ddPCR as well as studied by H&E staining and immunostaining for neuronal marker NeuN and for MTOR hyperactivation marker pS6 (**Fig. 5** and **Supplementary Table 6**). Notably, patient LR18-024 presented the *PIK3CA* p.E545K mutation at a very low level (VAF 0.28%) in only one (inferior temporal 4, IT4) of 9 fresh frozen brain samples, despite lack of clear dysplastic neuropathology in this region (**Fig. 5**, panel A’, see **Supplementary Table 6**). Conversely, the superior temporal region (fresh frozen sample ST2) that tested negative for the *PIK3CA* mutation by ddPCR demonstrated strongly positive pS6 staining and a dysplastic focus with clear FCD 2a neuropathology. Due to clinical and procedural workflows, the brain samples that were analyzed for neuropathology were FFPE blocks, while DNA analyzed initially for ddPCR was extracted from fresh frozen sections obtained from the same or contiguous areas, thus the molecular analysis was performed initially might represent a perilesional sublocations within the same resected brain sample. Therefore, to confirm these ultra-low level VAF and discern these correlations further in this patient, we obtained FFPE scrolls from these two regions (inferior and superior temporal) using the same blocks that were analyzed by neuropathology. For the inferior temporal, 3 blocks (IT 1, 2 and 3) were available and 20 sections each were collected. DNA from these three specimens were screened for all 6 hotspots, and only one of the three blocks (block IT2) had a positive result via ddPCR for *PIK3CA* p.E545K (VAF 0.14%), confirming that this area was indeed positive albeit at very low VAF%. For the superior temporal specimen, we were also able to perform manual dissection of the dysplastic area and non-dysplastic area (block ST1), comparing these with the full scrolls of the same block. Despite enriching for the affected area in this sample, we confirmed the absence of the *PIK3CA* p.E545K mutation, validating the results obtained from the fresh frozen sample (**Fig. 5, Supplementary Table 6**). A list of ddPCR results for the fresh frozen and FFPE samples for the inferior temporal and superior temporal specimens for individual LR18-024 is listed in **Supplementary Table 6**. The same was true for other cases, such as LR13-129 and LR16-313, where dysplastic and non-dysplastic areas are present at different levels in the same brain regions that tested positive via ddPCR (*MTOR* p.S2215F VAF 0-0.57% and *AKT3* p.E17K VAF 1.26-8.25%, respectively), with the level of dysplasia not matching the VAF% (**Fig. 5** panels A’’ and A’’’**, Supplementary Table 6**).

Finally, we compared the number of epilepsy surgeries and number of samples obtained per individual between hotspot mutation positive (*n*=17) and mutation negative cases (*n*=41). We found no statistically significant differences in the number of surgeries in the two populations; however, we found that the number of samples per patient obtained from mutation positive cases was significantly higher, with an average of 4.74 samples vs. 1.65 (*p-*value<0.01), compared to mutation-negative cases (**Supplementary Fig. 3**).

### Comparison of ddPCR with other molecular diagnostic methods

We compared ddPCR with two ultra-deep and highly targeted sequencing methods that have been used by us and others to detect mosaic mutations and accurately quantify the VAF in different tissues, including single molecule Molecular Inversion Probes (smMIPs), Amplicon Sequencing and exome sequencing.^26–30^ We compared results obtained overtime for 3 individuals (LR11-443, LR13-389 and LR16-004) (**Table 2**). While the level of concordance in the detection rate between Amplicon Sequencing and smMIPs was 100% (nine out of nine samples confirmed to be either positive or negative with both methods), the concordance level decreased when comparing ddPCR and smMIPs (13/16, 81.25%) or ddPCR and Amplicon Sequencing (6/9, 66.67%), due to presence of false positives (1 sample for case LR13-389, in red in **Table 2**) and false negatives (2 samples for case LR13-389 in blue in **Table 2**) in both smMIPs and Amplicon Sequencing results. Importantly, smMIPs and amplicon sequencing failed to detect several of the mutations detected by ddPCR due to sub-optimal depth of coverage of the former two methods compared to ddPCR which has the highest sensitivity among these methods.

**Table 2:**
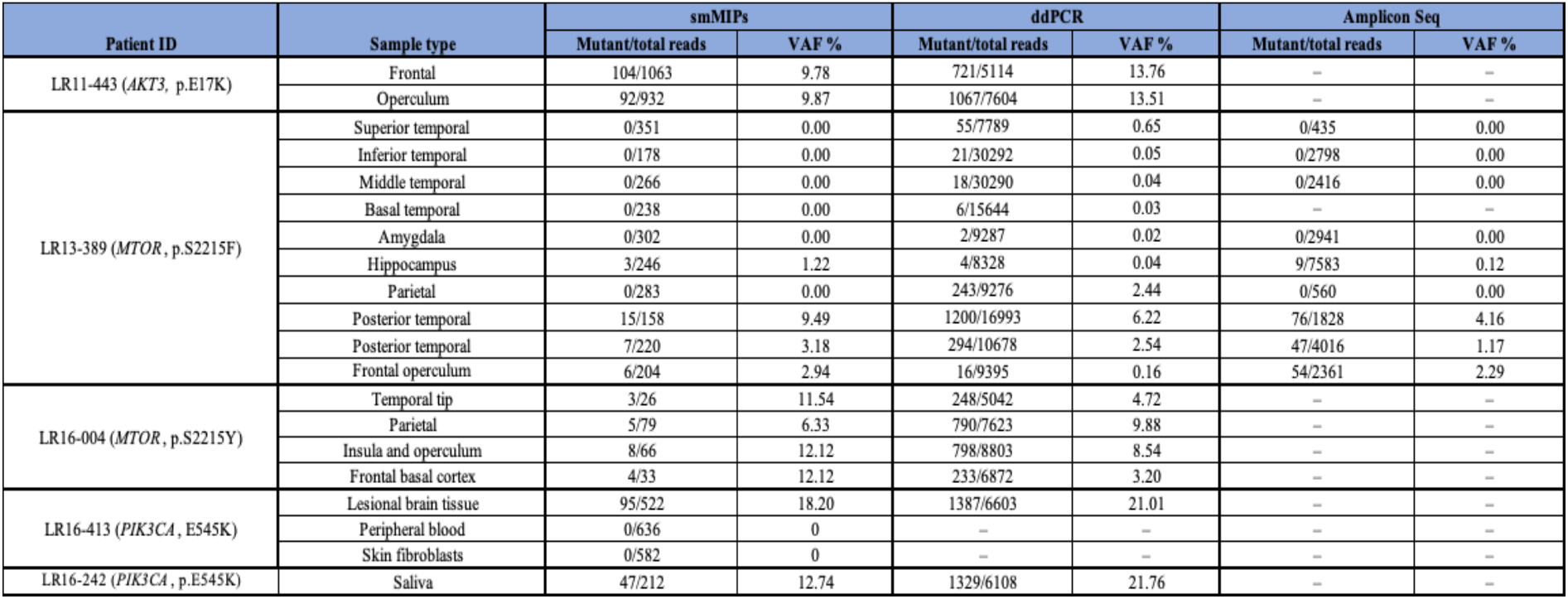
Comparison of ddPCR, smMIPs and Amplicon Sequencing for selected patients and brain samples.

## Discussion

In this study, we utilized an ultra-deep and highly targeted method to delineate the molecular spectrum of FMCD phenotypes caused by common or hotspot PI3K-AKT-MTOR pathway mutations. As recently reported by us and other groups, we confirmed a trend of association between FCD with *MTOR* hotspot mutations, as well as HMEG/DMEG and *PIK3CA* mutations.^9, 10^ In fact, among the FCD patients that were positive for a hotspot mutation, the two common *MTOR* variants were present in five cases (four cases positive for p.S2215F and one positive for p.S2215Y). In contrast, of our HMEG/DMEG cohort, 5 cases had the *PIK3CA* p.E545K variant, and one had p.E542K. *AKT3* p.E17K was also detected in both DMEG/HMEG and FCD cohorts (3/9 positive DMEG/HMEG patients; 1/7 positive FCD patients). Notably, there were differences in the molecular diagnostic yield between the subtypes of FCD, with only 1/10 of FCD 1 cases positive for one of the hotspot mutations, (10% of FCD 1 cohort, 33% of FCD 1b cohort); while 41.66% of our FCD 2 cohort had one of the hotspot mutations (10/24 cases), including specifically 42.10% of FCD 2a (8/19 individuals) and 50% of the FCD 2b (2/4). However, it is important to note that FCD 2a is the most common form of FCD overall and was present at higher frequency in our cohort (19 of the 40 individuals diagnosed with FCD by neuropathology, **Fig. 3B**). These differences support a possibly divergent genetic origin for FCD 1 and FCD 2, as suggested by us and other studies previously.^31^ Our study included only one individual with FCD 3 (LR17-349) who did not test positive for the six PI3K-AKT-MTOR mutations; however we only collected a saliva sample from this child. Our study further supported the evidence that the highly recurrent *AKT3* E17K variant is causal of HMEG and FCD with three children (LR11-443, LR13-351, LR16-313) all diagnosed with FCD 2a neuropathology. Unfortunately, neuropathologic evaluation of the fourth individual with the *AKT3* variant (LR12-317) was not available.

A key question in the field is whether there is a relationship between mutation burden in the brain and severity of cortical dysplasia, as there have several reports suggesting that there is no direct correlation between levels of mosaicism (or VAF) and phenotype severity.^19, 20, 32^ Therefore, we assessed whether there are any correlations between seizure age of onset and the VAF%, with the hypothesis that children with a higher mutation burden have earlier seizure onset, as a higher VAF might indicate an earlier developmental timepoint for the occurrence of the mutation in the developing brain with consequently major functional disruption of neuronal networks. In our cohort, all children with an average VAF over 5% had seizures within the first few months of life while individuals with VAF average lower than 1% had a delayed seizure onset. Our data therefore suggest that higher VAF% for these six hotspot mutations correlate with an earlier onset of epilepsy, and specifically that VAF >5% will lead to onset of seizures between day 0 and 30 of life, while VAF <1% tend to have later onset (3-6 years old). These findings may have a direct impact on prognosis and furthermore future eligibility for clinical trials using PI3K-AKT-MTOR pathway inhibitors. Similar genotype-phenotype correlations in *PIK3CA*-related vascular abnormalities were recently reported in a cohort of individuals.^13^ However, our data also demonstrate that there is no correlation between VAF and the severity of the brain malformation or structural lesions at the macroscopic (MRI) or microscopic (neuropathology) levels, as VAF% did not correlate directly with the extent of the cortical lesion or degree of dysplasia by imaging, confirming our findings and those of other groups.^8–10, 19, 32–35^ Retrospectively, we couldn’t delineate specific MRI findings that were exclusively present in the mutation positive cases and that would distinguish them from the negative ones. All in all, when comparing the two populations in our cohort (mutation positive vs mutation negative, **Supplementary Fig. 4**), the number of samples obtained per individual were different with many more samples obtained from mutation-positive individuals suggesting that perhaps more extensive resections have been performed in this cohort.

In our study, five individuals (LR12-317, LR13-129, LR15-251, LR16-313 and LR18-024) had a hotspot mutation detected in FFPE brain samples (**Supplementary Table 2**). For individual LR18-024, we were able to directly compare ddPCR results from fresh frozen and FFPE samples from the same brain region obtained during the same surgical procedure (**Fig. 5, Supplementary Table 6**). Not surprisingly, the VAF% was lower in the FFPE sample than the fresh frozen sample, possibly due to the increased DNA degradation and lower yeld in these samples. However, the variance might be due to an actual difference in the number of mutant cells in the two samples rather than to the method of sample preservation, as these two samples were from slightly different sub-regions, as discussed. For individual LR18-024 we were able to assess VAF on the same blocks that were used for neuropathology evaluation, and confirmed that the brain region that did not present dyplasia (inferior temporal) was positive for the *PIK3CA* p.E454K via ddPCR, while the brain region that did present a dysplastic focus (Superior temporal) was negative for this variant (**Fig. 5**, **Supplementary Table 6**).

Our study also demonstrated the utility of a ddPCR-based molecular diagnostic screen for mosaic mutations in suspected PI3K-AKT-MTOR related brain malformations (FCD, MEG and HMEG). Due to the nature of this spectrum and the implications of a molecular diagnosis in successfully treating these disorders, we believe that a first tier ddPCR-based molecular screen holds promise for improving the accuracy of detection of mutations. In highly mosaic disorders such as FCD and HMEG, pathogenic mutations can be present at extremely low levels, even in directly affected tissue.^1–6, 8, 10, 12, 19, 33^ For this reason, it is important to use a method with sufficient sensitivity to detect ultra low-level mosaicism; and our study shows that ddPCR provides an optimal approach for this purpose.^12, 14, 15^ When considering sample availability, it is very imperative to select the relevant tissue to efficiently identify mutations. We were able to demonstrate the importance of the number of samples and types of samples obtained per individual to identify a causal mutation. We obtained a better solve rate when testing at least 2 brain samples per individual (solve rate 53.85%, **Fig. 2**). However, brain samples might not be available for children who are not eligible for epilepsy surgery, thus a second tissue source could be saliva, as we were able to detect hotspots mutations in 6 out of seven positive cases, with only one positive individual (LR13-389) having a negative result in saliva while presenting a VAF of 1.66-6.22% in the brain (cohort saliva solve rate 25%, **Supplementary Fig. 1**). We were not able to detect mutations in blood or skin, although these latter samples were the minority in our cohort (*n*=5). While ddPCR provides highly targeted ultra-high depth coverage, it is a low-throughput method, with probes designed to detect specific mutations. Therefore, ddPCR is inefficient in disorders with a wide mutational spectrum. We acknowledge that the molecular spectrum of these disorders is constantly expanding and while the majority of genetic variants are mosaic, a fraction of individuals have germline or constitutional variants in other genes such as *DEPDC5* or *SLC35A2*, among others.^19, 36, 37^ Table 1 summarizes reported cases of FCD, HMEG, or DMEG caused by pathogenic mutations in *PIK3CA*, *AKT3*, or *MTOR*. We molecular solved 29.31% of individuals in this series. Importantly, we were able to solve 9 of the 11 cases with HMEG/DMEG (81.81%) and 10 of the 24 cases diagnosed with FCD 2a, highlighting the contribution of *PIK3CA* and *AKT3* hotspot mutations in HMEG/DMEG and *MTOR* mutations in FCD 2a. Overall, these data support that screening these common mutations comprises a possibly efficient first tier molecular diagnostic approach. The additional advantage of utilizing a ddPCR-based approach is the minimal input DNA requirement. Our assay used only used 8 ng of input genomic DNA per well, with four replicate wells per hotspot. This amount of DNA is generally available from most clinical samples, and this is a specific advantage with ddPCR compared to other deep sequencing techniques such as smMIPs, exome and genome sequencing which have much higher genomic input requirements while also having a lower sensitivity for the mosaicism detection. Our results show that ddPCR has deeper coverage when compared with these other methods using the same DNA input, and is able to discriminate type 1 and type 2 errors (false positives and false negatives), demonstrating higher specificity and sensitivity. Additionally, ddPCR testing is optimal for low-quality or degraded DNA, such as FFPE samples,^11^ and samples containing traces of substances known to inhibit regular PCR reactions.^38^

In conclusion, our study shows the correlation between the age of onset of the epileptic phenotype in FCD and DMEG/HMEG with mutational burden (mosaicism level). Furthermore, our results confirm the utility of ddPCR in a clinical setting for targeted testing of this under-characterized spectrum in order to successfully detect low-level pathogenic variants, allowing for early molecular diagnosis and facilitating recurrence risk counseling and, more importantly, molecularly targeted therapies (i.e., PI3K-AKT-MTOR pathway inhibitors).

## Data Availability

All data generated or analysed during this study are included in this published article and its
supplementary information files. The raw data generated during and/or analyzed during the
current study are available from the corresponding author on request.

## Acknowledgements

We thank the families and their provider for their participation in this study.

## Supplementary material

‘Supplementary material is available at *Brain* online’

---

**Supplementary Table 1:** ddPCR probe specifications. Commercially available probes were purchased from Biorad and used following manufacturer’s instructions for amplification cycles, with annealing temperatures as shown (in °C)

| Probeset description | Assay ID | Annealing temperature |
| --- | --- | --- |
| ddPCR™ Mutation Assay: AKT3 p.E17K, Human | dHsaMDS121759946 | 55 |
| ddPCR™ Mutation Assay: PIK3CA p.E542K, Human | dHsaMDV2010073 | 57 |
| ddPCR™ Mutation Assay: PIK3CA p.E545K, Human | dHsaMDV2010075 | 57 |
| ddPCR™ Mutation Assay: PIK3CA p.H1047R, Human | dHsaMDV2010077 | 55 |
| ddPCR™ Mutation Assay: MTOR p.S2215F, Human | dHsaMDS365202467 | 55 |
| ddPCR™ Mutation Assay: MTOR p.S2215Y, Human | dHsaMDS2514998 | 55 |

**Supplementary Table 2:** Complete list of samples and ddPCR results for the 17 mutation positive patients. Patients’ de-identified code (Patient-ID), diagnosis based on neuroimaging (Dx-MRI), Diagnosis based on neuropathology (Dx-NPath), hotspot variant, sample type and ddPCR results (Variant allele fraction [VAF%], Delta confidence interval [DCI], and number of wild-type and mutant droplets [average of quadruplicate]) are listed for each of the 17 mutation-positive patients. Samples were considered positive for a given mutation when DCI>0.045 and VAF>0.05% (shaded in green). Number of wild-type droplets is indicative of ddPCR run quality: samples with >3000 droplets are shaded in green. Samples with <3000 droplets (shaded in yellow) were retained only if the samples had a DCI>1% and/or other samples of the same patients had clear positive results.

| Patient ID | Dx - MRI | Dx - NPATH | Genetic variant | Sample type | VAF% | DCI | WT droplets | Mutant droplets |
| --- | --- | --- | --- | --- | --- | --- | --- | --- |
| LR12-246 | HMEG | FCD 2a | *MTOR* (p.S2215F) | Brain | 0.16 | 0.047 | 10945 | 19 |
| LR13-129 | FCD | FCD 2b | *MTOR* (p.S2215F) | Brain, lateral front lobe | 0.02 | -0.051 | 10946 | 2 |
|  |  |  |  | Brain, medial front lobe | 0.04 | -0.043 | 12079 | 6 |
|  |  |  |  | Brain, orbital front lobe | 0.57 | 0.388 | 11908 | 75 |
|  |  |  |  | Brain, FFPE | 0.06 | -0.034 | 12784 | 8 |
|  |  |  |  | Brain, FFPE | 0.05 | -0.047 | 10156 | 5 |
|  |  |  |  | Brain, FFPE | 0.24 | 0.104 | 11220 | 30 |
|  |  |  |  | Brain, FFPE | 0.03 | -0.051 | 11116 | 4 |
|  |  |  |  | Brain, FFPE | 0.06 | -0.040 | 8708 | 6 |
|  |  |  |  | Brain, FFPE | 0.08 | -0.025 | 10519 | 9 |
| LR13-389 | FCD | FCD 2a | *MTOR* (p.S2215F) | Brain, superior temporal | 0.65 | 0.390 | 7734 | 55 |
|  |  |  |  | Brain, inferior temporal | 0.05 | -0.030 | 30271 |  |
|  |  |  |  | Brain, middle temporal | 0.04 | -0.030 | 30272 |  |
|  |  |  |  | Brain, basal temporal | 0.03 | -0.050 | 15638 |  |
|  |  |  |  | Brain, amygdala | 0.02 | -0.047 | 9285 | 2 |
|  |  |  |  | Brain, hippocampus | 0.04 | -0.047 | 8324 | 4 |
|  |  |  |  | Brain, parietal lobe | 2.44 | 2.090 | 9033 | 243 |
|  |  |  |  | Brain, posterior | 6.22 | 5.824 | 15793 | 1200 |
|  |  |  |  | Brain, posterior | 2.54 | 2.202 | 10384 | 294 |
|  |  |  |  | Brain, frontal | 0.16 | 0.032 | 9379 | 16 |
|  |  |  |  | Saliva | 0.03 | -0.047 | 7165 | 2 |
|  |  |  |  | Brain, mid-frontal | 0.05 | -0.047 | 8199 | 4 |
|  |  |  |  | Brain, posterior frontal | 4.16 | 3.727 | 8990 | 421 |
|  |  |  |  | Brain, posterior | 1.66 | 1.346 | 7949 | 143 |
|  |  |  |  | Brain, inferior frontal | 0.10 | -0.012 | 8087 | 9 |
|  |  |  |  | Brain, orbital frontal | 0.11 | -0.010 | 7522 | 9 |
| LR14-046 | FCD | FCD | *MTOR* (p.S2215F) | Brain, deep temporal | 3.91 | 3.379 | 7315 | 323 |
| LR14-155 | FCD | FCD 2b | *MTOR* (p.S2215F) | Brain, posterior frontal | 0.34 | 0.121 | 10149 | 38 |
|  |  |  |  | Brain, anterior frontal | 0.06 | -0.091 | 11304 | 8 |
| LR13-197 | DMEG | FCD 1b | *PIK3CA* (p.E545K) | Blood | 0.04 | -0.077 | 9276 | 4 |
|  |  |  |  | Saliva | 20.68 | 19.398 | 2984 | 792 |
| LR15-251 | HMEG | NA | *PIK3CA* (p.E545K) | Brain, FFPE | 18.86 | 16.691 | 1051 | 246 |
|  |  |  |  | Blood | 0.02 | -0.049 | 11067 | 2 |
|  |  |  |  | Buccal | 0.12 | -0.015 | 6464 | 8 |
| LR16-242 | HMEG | NA | *PIK3CA* (p.E545K) | Saliva | 21.76 | 20.780 | 6108 | 1757 |
| LR16-413 | HMEG | FCD | *PIK3CA* (p.E545K) | Brain | 21.01 | 19.996 | 6603 | 1827 |
|  |  |  |  | Saliva | 22.67 | 21.459 | 4560 | 1377 |
| LR18-024 | FCD | FCD 2a | *PIK3CA* (p.E545K) | Brain, anterior dysplasia | 0.03 | -0.072 | 9793 | 3 |
|  |  |  |  | Brain, superior temporal | 0.06 | -0.050 | 14216 | 10 |
|  |  |  |  | Brain, temporal tip | 0.13 | 0.000 | 11545 | 17 |
|  |  |  |  | Brain, inferior temporal | 0.28 | 0.120 | 14078 | 44 |
|  |  |  |  | Brain, anterior inferior | 0.15 | 0.010 | 12641 | 21 |
|  |  |  |  | Brain, middle temporal | 0.10 | -0.040 | 9034 | 10 |
|  |  |  |  | Brain, amygdala | 0.12 | 0.000 | 15147 | 21 |
|  |  |  |  | Brain, hippocampus | 0.11 | -0.050 | 6252 | 7 |
|  |  |  |  | Brain, cusa | 0.05 | -0.072 | 4266 | 2 |
| LR11-443 | HMEG | FCD 2a | *AKT3* (p.E17K) | Brain, frontal lobe | 13.76 | 12.771 | 4393 | 721 |
|  |  |  |  | Brain | 13.51 | 12.703 | 6537 | 1067 |
| LR12-317 | HMEG | NA | *AKT3* (p.E17K) | Saliva | 0.58 | 0.208 | 2333 | 14 |
|  |  |  |  | Brain, FFPE | 12.93 | 10.461 | 663 | 99 |
| LR13-351 | FCD | FCD 2a | *AKT3* (p.E17K) | Brain | 0.62 | 0.428 | 7902 | 53 |
|  |  |  |  | Blood | 0.03 | -0.023 | 9090 | 3 |
| LR16-313 | FCD | FCD 2a | *AKT3* (p.E17K) | Skin | 0.00 | 0.000 | 3688 | 0 |
|  |  |  |  | Brain, temporal lob | 1.66 | 1.231 | 3206 | 57 |
|  |  |  |  | Brain, hippocampus | 1.26 | 0.639 | 1238 | 16 |
|  |  |  |  | Brain, insula | 7.26 | 6.384 | 3017 | 245 |
|  |  |  |  | Brain, insula | 5.93 | 5.395 | 6603 | 453 |
|  |  |  |  | Brain, temporal tip | 2.13 | 1.670 | 3538 | 80 |
|  |  |  |  | brain, FFPE | 6.44 | 5.315 | 1914 | 134 |
|  |  |  |  | brain, FFPE | 7.08 | 6.033 | 2432 | 189 |
|  |  |  |  | brain, FFPE | 8.25 | 6.990 | 1849 | 169 |
|  |  |  |  | brain, FFPE | 2.44 | 1.608 | 1537 | 39 |
| LR16-004 | HMEG | FCD 2a | *MTOR* (p.S2215Y) | Saliva | 0.96 | 0.761 | 8906 | 92 |
|  |  |  |  | Brain, lateral temporal | 7.18 | 6.626 | 7365 | 601 |
|  |  |  |  | Brain, inferior temporal | 1.05 | 0.836 | 7865 | 89 |
|  |  |  |  | Brain, temporal tip | 4.72 | 4.143 | 4794 | 248 |
|  |  |  |  | Brain, hippocampus | 0.03 | 0.000 | 6075 | 2 |
|  |  |  |  | Brain, anterior frontal | 0.02 | 0.000 | 6008 | 1 |
|  |  |  |  | Brain, middle frontal | 13.59 | 12.756 | 5361 | 877 |
|  |  |  |  | Brain, parietal lobe | 9.88 | 9.224 | 6833 | 790 |
|  |  |  |  | Brain, insula and | 8.54 | 7.973 | 8005 | 798 |
|  |  |  |  | Brain, ventricle wall | 1.60 | 1.341 | 8404 | 147 |
|  |  |  |  | Brain, frontal basal | 3.20 | 2.798 | 6639 | 233 |
|  |  |  |  | Brain, gyrus rectus | 0.00 | 0.000 | 8727 | 0 |
| LR16-470 | HMEG | NA | *PIK3CA* (p.E542K) | Saliva | 1.47 | 1.056 | 4498 | 70 |
| LR12-251 | FCD | FCD 2a | *PIK3CA* (p.H1047R) | Brain | 4.58 | 4.186 | 10877 | 574 |

**Supplementary Table 3:** Summary of clinical and neuroimaging features for the 17 mutation positive patients. Abbreviations: d, day; DMEG, dysplastic megalencephaly; FCD focal cortical dysplasia; HMEG, hemimegalencephaly; y, year; m, month; PQD, posterior quadrant dysplasia; N/A, not available. Data from two patients have been previously published, namely: LR13-389 (PMID 27159400); LR11-443 (PMID 28969385, 25722288, 27159400).

| Mutation | Patient ID | Diagnosis | Head size (OFC) at birth | Head size (OFC) last assessed | Seizure – age at onset | Seizure – types | Seizure – medical Rx | Brain MRI – ages | Brain MRI – findings | Epilepsy surgery – types | Neuropathology | Post surgery – epilepsy recurrence | Somatic findings (vascular malformations, overgrowth) |
| --- | --- | --- | --- | --- | --- | --- | --- | --- | --- | --- | --- | --- | --- |
| MTOR<br>p.S2215F | LR12-246 | HMEG (PQD) | 38.5cm at 1m | 43.80cm | 3d | Multiple types | Medically intractable | 6m | PQD | Hemispherectomy (x2) | FCD2a | No postsurgical recurrence | – |
|  | LR13-129 | FCD | N/A | 49cm | 11m | drop attacks, gaze seizures, intractable | Medically intractable | 5y | FCD | Craniotomy NSR (3), craniotomy hemispherectomy | FCD2b (x3, not seen in last resection) | Possible 1 seizure recurrence | – |
|  | LR13-389 | FCD | 37.1cm at 6w | 48.2cm at 3y | 5w | CPS | Medically intractable | 7m | HMEG | Craniotomy NSR (12/30/13), craniotomy hemispherectomy (5/22/14) | FCD2a | Only rare seizures of a different semiology post 2nd procedure | – |
|  | LR14-046 | FCD | N/A | 53.4cm | 7m | Staring spells with occasional/AI left upper extremity twitching, tonic posturing | Medically intractable | 5y | FCD | Craniotomy placement electrode, craniotomy removal grid w/ resection | FCD2b | No postsurgical recurrence | – |
|  | LR14-155 | FCD | N/A | 53.5cm (12y) | 5y | Localization-related (focal) epilepsy, CPS | Medically intractable | 12y, 15y | FCD | Craniotomy NSR | FCD2b | No postsurgical recurrence | – |
| PIK3CA<br>p.E545K | LR13-197 | DMEG | N/A | 38cm | ~29d | Staring episodes, back arching, focal | Medically intractable | 37d | HMEG | N/A | N/A | N/A | Right lower extremity hemihypertrophy with some left-sided facial asymmetry |
|  | LR15-251 | HMEG | N/A | N/A (WNL by report) | 1d | Status epilepticus; tonic-clonic with breath arrest | Currently well-controlled, last seizure 4 years ago | 3y | HMEG | Temporo-parietal-occipital functional disconnection (2005), intracranial stereotaxis (May 2008), temporo-parietal-occipital functional disconnection, more substantial than 1st procedure (Sept 2008) | N/A | N/A | Flat feet, translucent skin, sebaceous nevus on left cheek |
|  | LR16-242 | HMEG | 37.2 cm | 52 cm at 3y7m, 95% for age | 1 d | Flexor spasms, complex partial seizures | Medically intractable, much improved on ketogenic diet | 2d | R HMEG, with L hemimicrocephaly | No | N/A | N/A (died at age 4y1m from respiratory arrest after status epilepticus) | Enlarged R cheek and eye, swirled brown macule on R cheek extending to neck |
|  | LR16-413 | HMEG | 40cm | N/A | 2d | Abnormal eye movements, TC, EEG c/w Ohtahara syndrome | Medically intractable | N/A | L HMEG and R perisylvian FCD (likely FCD type 2) | N/A | N/A | N/A | Hypertrophic left lip/cheek |
|  | LR18-024 | FCD | N/A | N/A | 6y | Abnormal eye movements, complex partial | Medically intractable | 5y, 15y, 17y | Nonspecific small focal areas of T2/FLAIR prolongation in the L superior frontal periventricular white matter, hypometabolism in R anterior temporal region | Microdissection | FCD2a | No postsurgical recurrence | – |
| AKT3<br>p.E17K | LR11-443 | HMEG (+ contralateral focus) | 38cm | 48.4cm | 3d | Generalized, focal | Medically intractable, ketogenic diet | 4y | HMEG with involvement of contralateral mesial occipital lobe | Craniotomy hemispherectomy total, craniotomy | FCD2a | Postsurgical recurrence a few days postop #1, and after ~6m postop#2 | Cutis marmorata, organomegaly, L retroorbital vascular malformation |
|  | LR12-317 | HMEG | N/A | 44.3 | 7d | [stiffness, flexion of trunk, rhythmic lip smacking/chewing, rigid extremities which moved from flexion into extension 3 times] | Medically intractable | 13d | HMEG | Hemispherectomy | [Sections show disorganized cortical layering; neurons disorganized with focal clustering and vary immensely in size, polarity inconsistently maintained, Dx of HMEG, MCAP, Ohtahara syndrome] | No postsurgical recurrence | Cutaneous vascular lesions on extremities and trunk |
|  | LR13-351 | FCD | N/A | N/A | N/A | CPS | N/A | N/A | N/A | Right occipital lobectomy | FCD2a | Rare seizures | – |
|  | LR16-313 | FCD | 36.8 (1m) | 45.9 (2y) | 4d | ISS | Medically intractable | 1m | HMEG | Right craniotomy hemispherectomy | FCD2a | No postsurgical recurrence | – |
|  | LR16-004 | HMEG | N/A | 45.9 (3y) | 3w | ISS | Medically intractable | 6m | HMEG | Craniotomy, left hemispherectomy total (1/7/16) | FCD2a | Movements continued to be jerky after surgery, but no epilepsy recurrence | – |
| PIK3CA<br>p.E542K | LR16-470 | HMEG | N/A | N/A | N/A | Focal epileptic spasms | N/A | N/A | N/A | N/A | N/A | N/A | Abnormal swirling skin pigmentation on several areas of the body |
| PIK3CA<br>p.H1047R | LR12-251 | FCD | N/A, referred after 1st seizure | 50cm | 3y | Afebrile, partial seizures | Medically intractable | 5y | FCD with mass effect | R temporoparietal craniotomy, R occipital lobectomy | FCD2a | No postsurgical recurrence | – |

**Supplementary Table 4:**
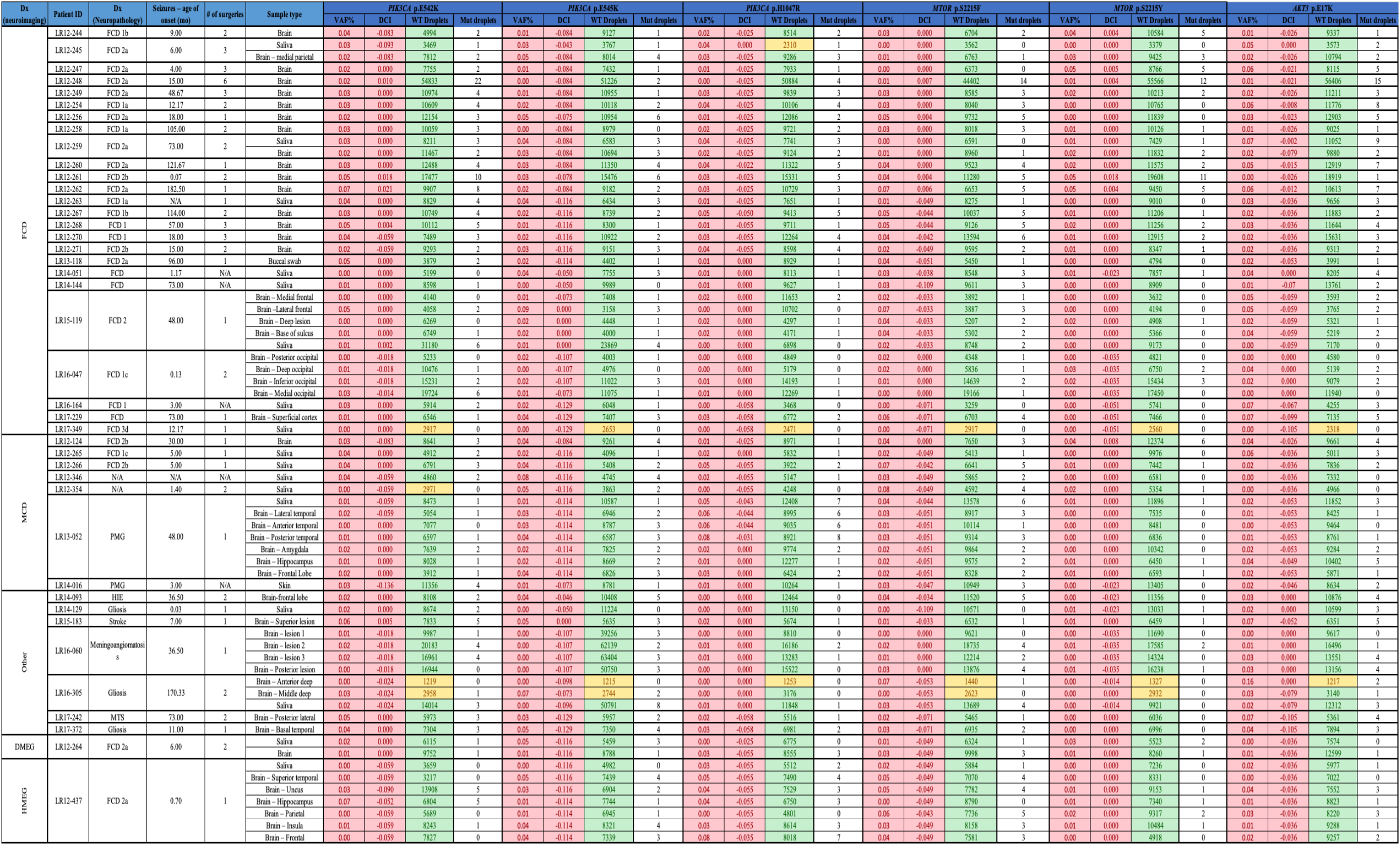
Data summary of patients and samples who tested negative for the 6 PI3K-AKT-MTOR hotspot mutations. Patients’ de-identified code (Patient-ID), diagnosis based on neuroimaging (Dx-MRI), Diagnosis based on neuropathology (Dx-NPath), and sample type are listed for the mutation-negative patients (n=41). All samples were tested via ddPCR for all 6 hotspots in quadruplicates and in multiple independent runs. All samples had a DCI<0.045, thus they were considered negative (cells shaded in red). The average of each quadruplicate is reported here. Number of WT droplets is reported as measure of quality of the ddPCR run; samples with over 3000 were considered robust and subsequently analyzed (green shaded cells). If samples had less than 3000 WT droplets (yellow shaded cells) and VAF<0.1%, they were deemed negative. When multiple runs were performed for a given sample, results from the run yielding the highest number of wild-type droplets (WT droplets) is reported here. Patient LR12-245 was previously published in PMID: 27159400.

**Supplementary Table 5:**
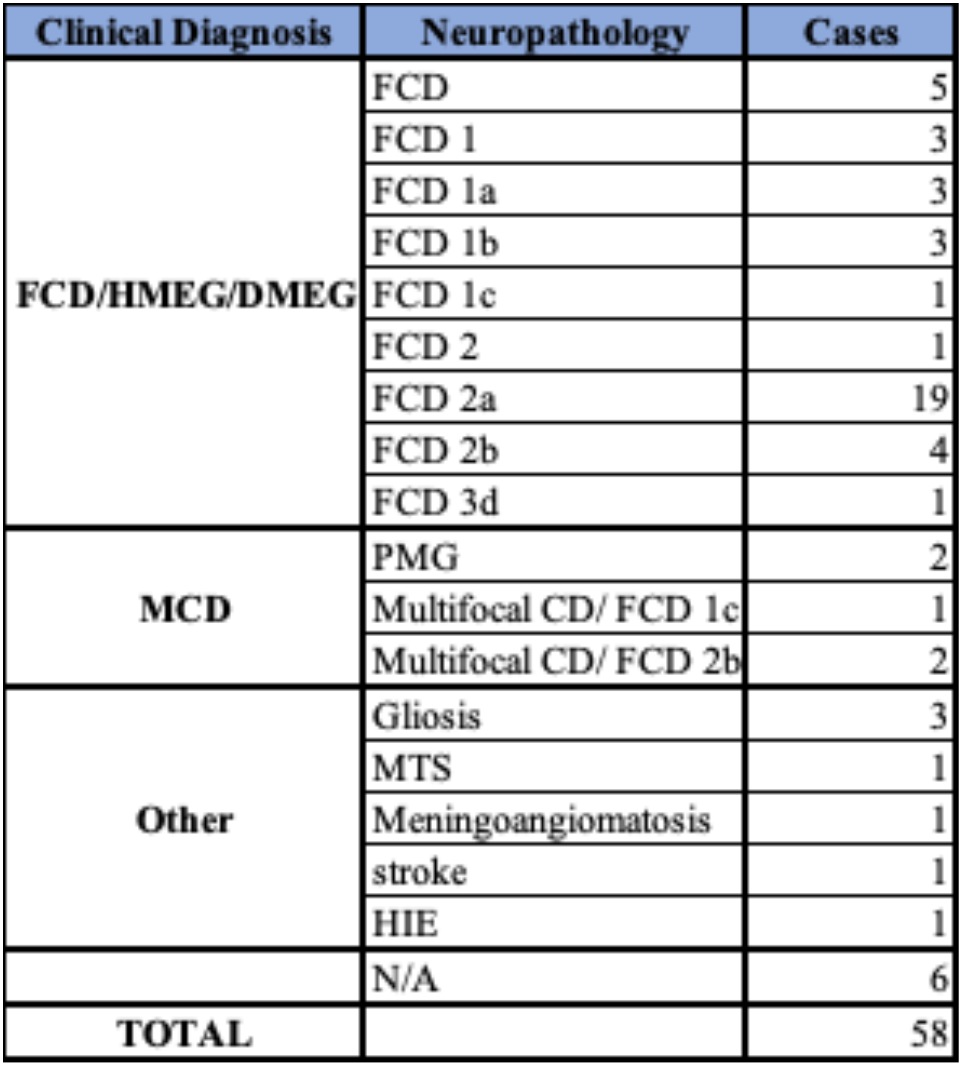
Neuropathological classification for cohort. The neuropathological diagnosis for the 58 patients reported here, sub-classified by their primary clinical diagnosis, and the number of cases within each category.

**Supplementary Table 6:**
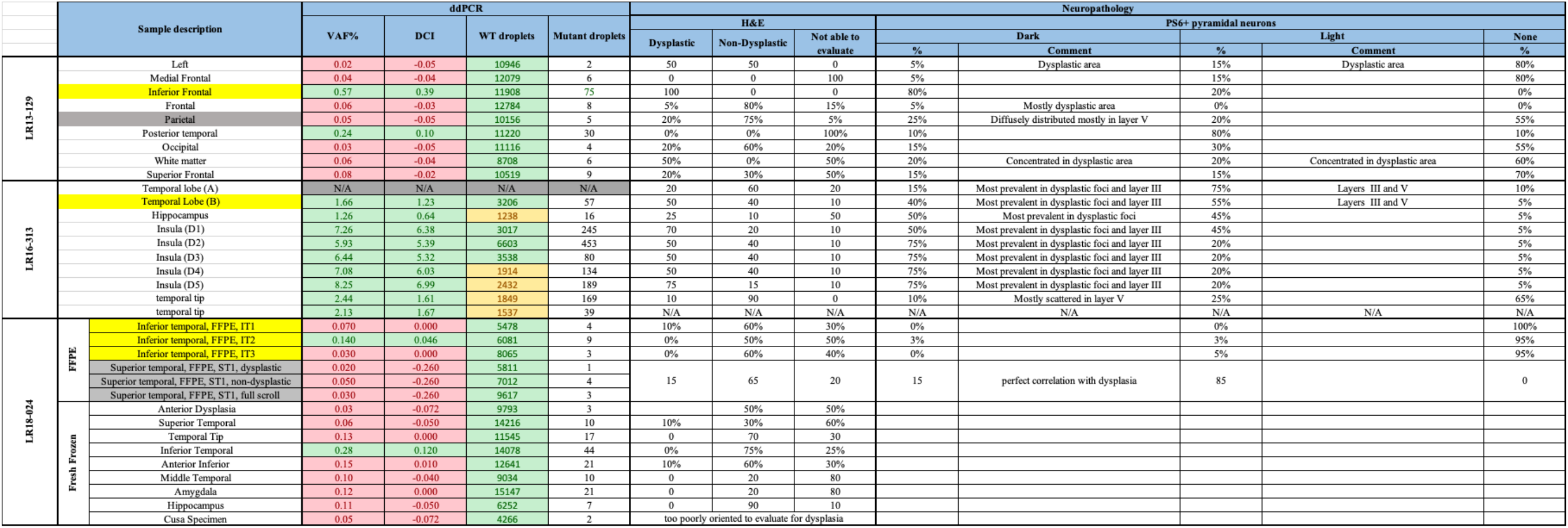
Genotype-neuropathology correlation for patient LR13-129; LR16-313 and LR18-024. Results from ddPCR analysis and neuropathology evaluation for these three representative patients are listed below. For neuropathologic evaluation, hematoxylin and eosin staining (H&E) was performed and cortical layering, as well as presence/absence of balloon/abnormal neurons was evaluated. The H&E results refer to the percentage of abnormal area as evaluated by the pathologist. When possible, pS6 staining was performed and presence of positive neurons (dark or light staining, percentage relative to the area of the section) was performed. For ddPCR results, cells shaded in gray represent brain regions that are positive for the relative hotspot mutation, while cells shaded in gray represent brain regions that do not present the mutation. For LR18-204, FFPE and fresh frozen samples are listed. Two representative brain regions (inferior temporal, positive; and superior temporal, negative) from patient LR18-024 were selected for a comparison of VAF% detected via ddPCR in fresh frozen vs FFPE samples. See Figure 5 for a schematic representation of the two LR18-024 specimens and relative neuropathology findings, and Supplementary Fig. 3 for neuropathology analysis for samples relative to LR13-129 and LR16-313. Blocks highlighted in Fig.5 and Supplementary Fig. 3 are highlighted here in yellow (positive blocks) and gray (negative blocks).

## SUPPLEMENTARY FIGURES

**Supplementary Figure 1.**
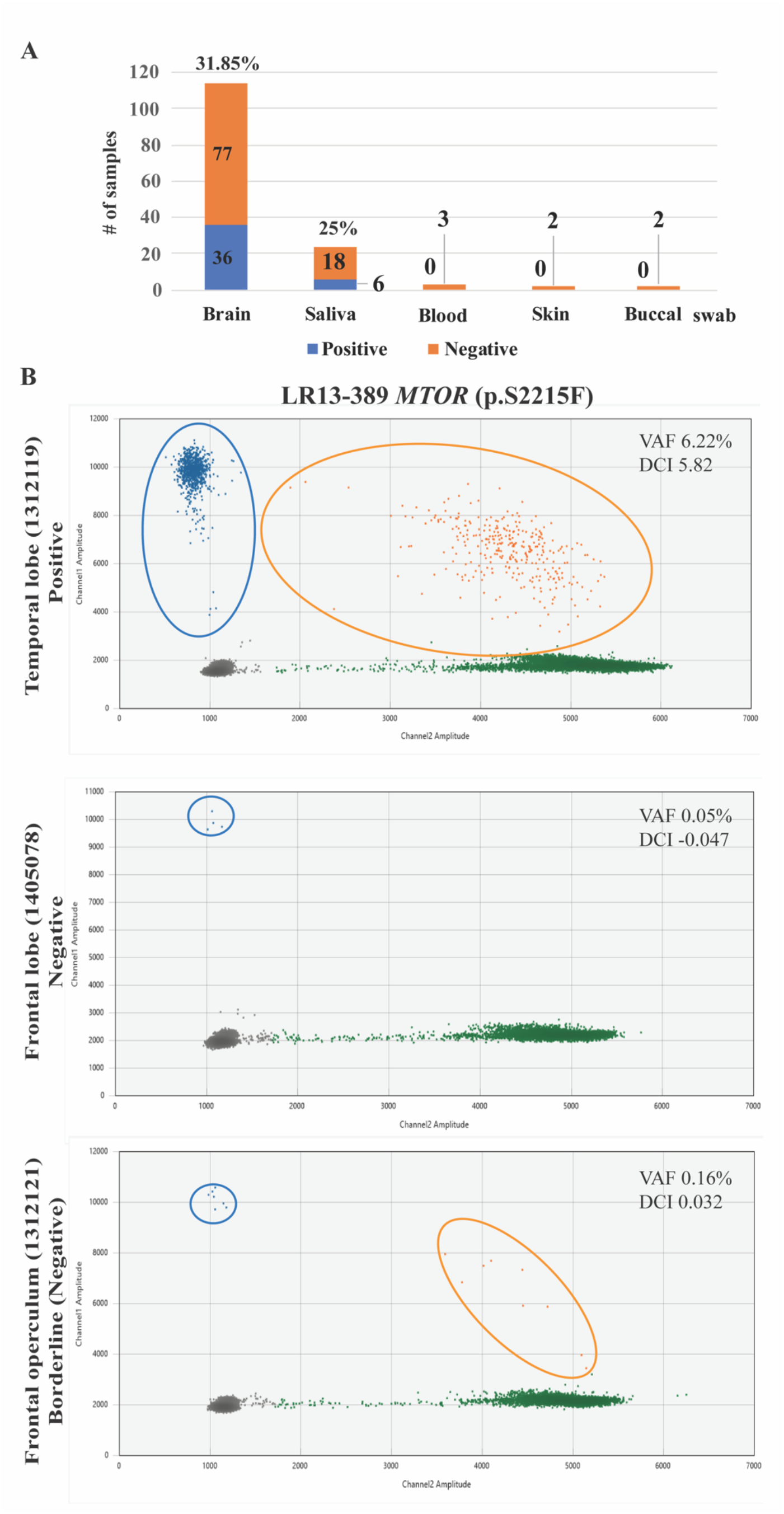
ddPCR representative results. **(A)** The solve rate per type of tissue based on number of positive specimens vs. total. Hotspot mutations were not detectable in blood, skin or buccal swab. **(B)** Examples of 2D Quantasoft plots for brain tissues from patient LR13-389, carrying the *MTOR* p.S2215F mutation. Variant allele fractions (VAF%) and Delta confidence intervals (DCI) are shown for each sample. The temporal lobe (sample 1312119) shows a clear positive result, with mutant droplets (in the blue oval) clustering in the upper left quadrant and in higher number than the double positive droplets (in the orange oval), with wild-type droplets in green clustering in the lower right quadrant. A frontal lobe sample from the second surgical procedure (sample 1405078) was negative, with 4 mutant droplets (in the blue oval) detected which were also present in the wild-type control in the same run, as demonstrated by the negative DCI. A sample from the frontal operculum region (1312121) had a “borderline” result. Samples with VAF 0.16% and DCI=0.032 would be considered positive by the Biorad analysis protocol. However, as the plot shows, the number of mutant droplets (in the blue oval, n=7) are fewer than the double positive droplets (in the orange oval, n=9). The high number of double positive droplets leads to a false positive result, thus samples with DCI<0.045 are considered negative in this study.

**Supplementary Figure 2:**
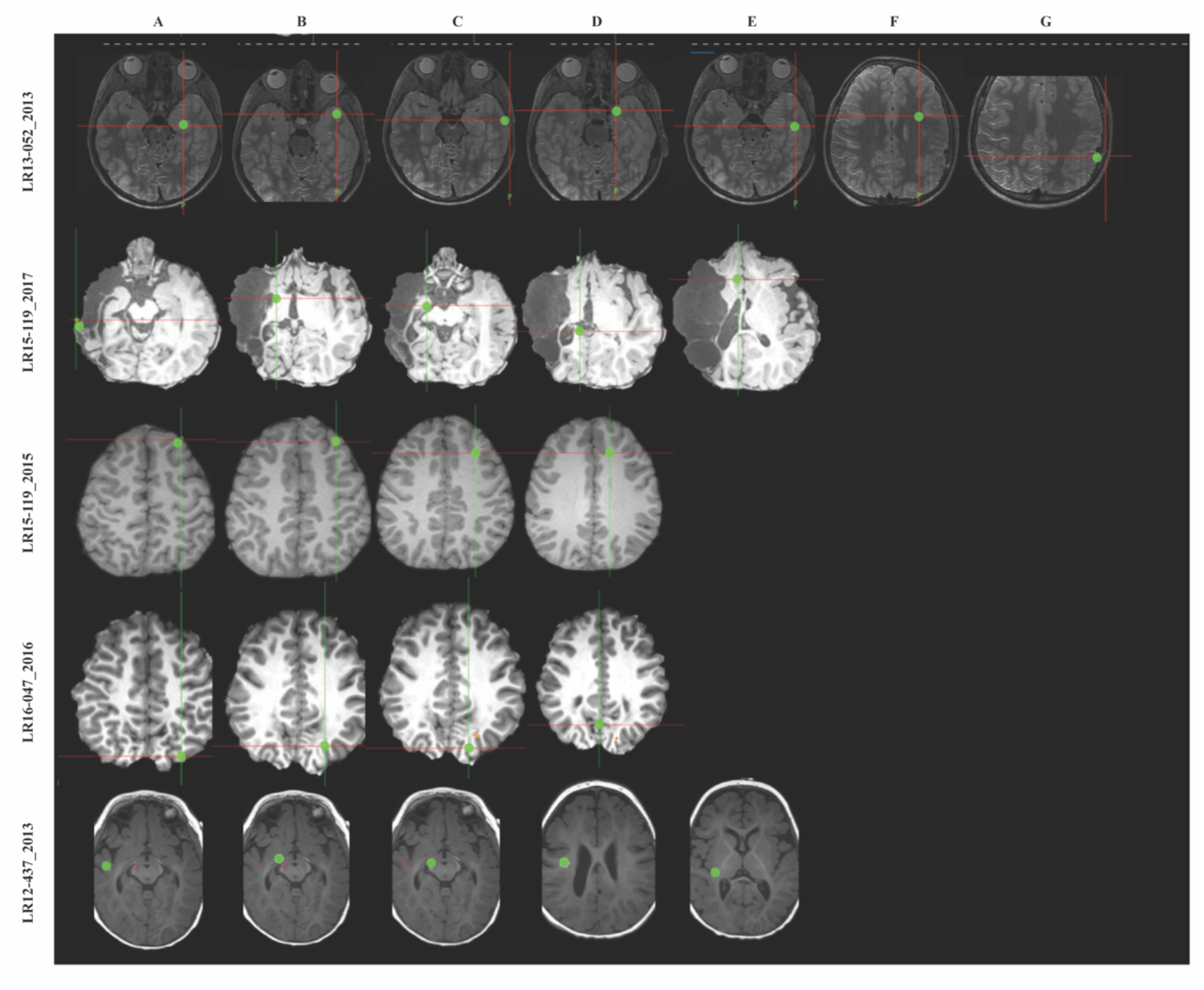
Brain MRI of four patients with FCD/HMEG/DMEG who tested negative for the 6 PI3K-AKT-MTOR hotspot mutations. Summary MRI images showing biopsy locations. For each image, cross-hairs reflect the sample site, with letters reflecting the multiple samples taken. Green dots represent the exact location of resection. The year in which the surgery was performed is indicated after the underscore, as patient LR15-119 underwent multiple brain surgeries. These samples were tested via ddPCR for the 6 hotspot mutations and were all negative.

**Supplementary Figure 3:**
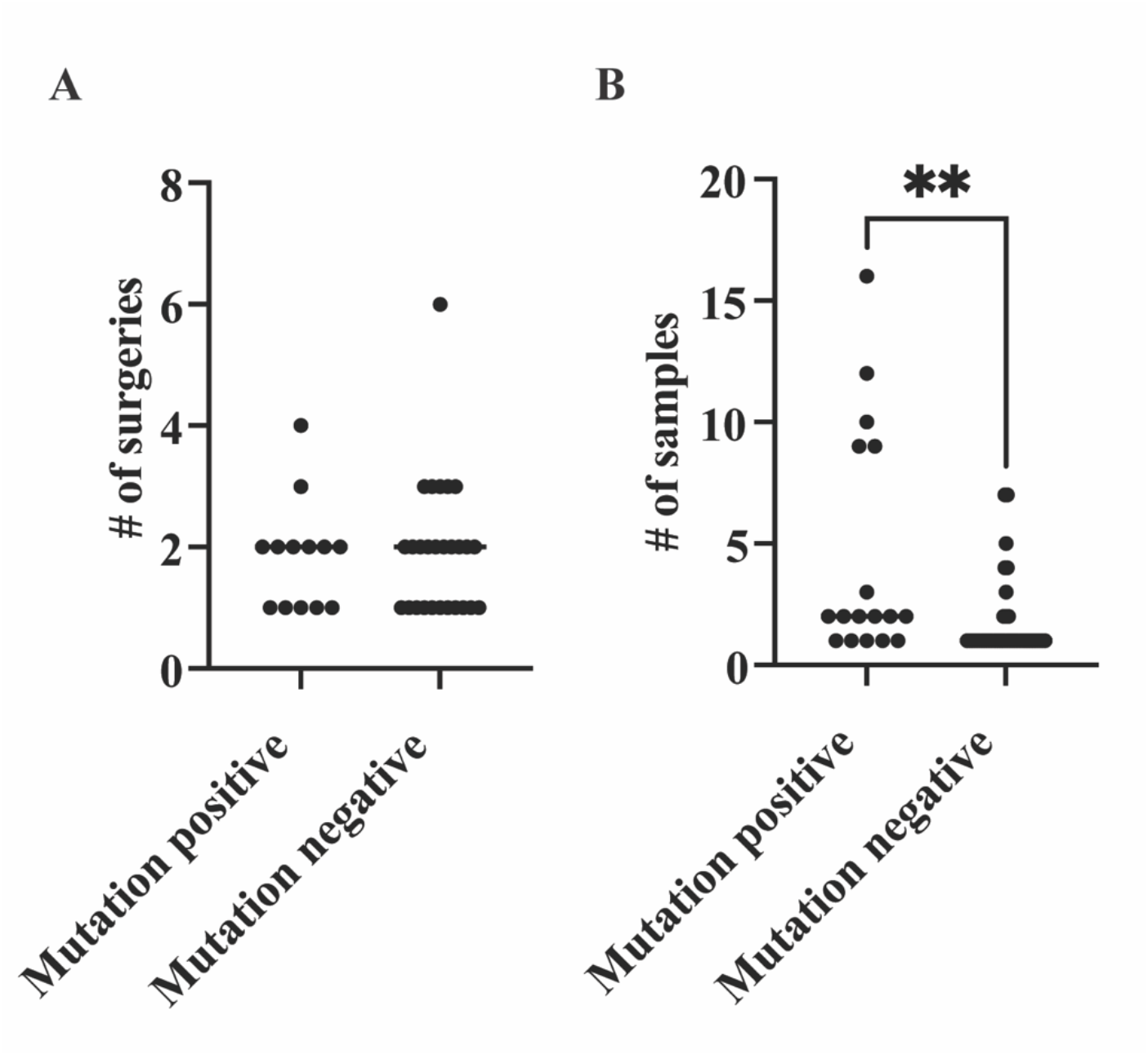
Comparison of number of surgeries and samples in the hotspot mutation positive and negative patients. **(A)** The number of surgeries per individual did not differ among the two populations. (**B)** The number of specimens per individual was significantly higher in the mutation-positive cohort (Kolmogorov-Smirnov test, *p*value=0.0068). Notably, our results do not rule out the possibility of a genetic mutation in the negative cases.

